# Developing and externally validating machine learning models to forecast short-term risk of ventilator-associated pneumonia

**DOI:** 10.64898/2026.01.28.26344858

**Authors:** Alec K. Peltekian, Wan-Ting Liao, Vijeeth Guggilla, Nikolay Markov, Karolina Senkow, Zewei Liao, Marjorie Kang, Luke Rasmussen, Elsa Tavernier, Stephan Ehrmann, Rebecca K. Clepp, Thomas Stoeger, Theresa Walunas, Alok Choudhary, Alexander V. Misharin, Benjamin D. Singer, GR Scott Budinger, Richard G. Wunderink, Catherine A. Gao, Ankit Agrawal, The NU SCRIPT Study Investigators

**Affiliations:** Department of Computer Science, Northwestern University McCormick School of Engineering and Applied Science, Chicago, IL, USA; Division of Pulmonary and Critical Care, Northwestern University Feinberg School of Medicine, Chicago, IL, USA; Division of Biostatistics and Informatics, Northwestern University Feinberg School of Medicine, Chicago, IL, USA; Division of Pulmonary and Critical Care, Department of Medicine, University of Chicago, Chicago, IL, USA; CIC, INSERM 1415, CHRU Tours, Tours, France; Methods in Patients-Centered Outcomes and Health Research SPHERE, INSERM 1246, Tours and Nantes, France; Médecine Intensive Réanimation, INSERM CIC1415, CRICS-TriggerSEP F-CRIN researche network, CHRU de Tours, Tours, France; Centre d’étude des pathologies respiratoires, INSERM U1100, Université de Tours, Tours, France; The Potocsnak Longevity Institute, Northwestern University, Chicago, IL, USA; Simpson Querrey Lung Institute for Translational Science, Northwestern University Feinberg School of Medicine, Chicago, IL, USA; NSF-Simons National Institute for Theory and Mathematics in Biology, Chicago, IL, USA; Department of Electrical and Computer Engineering, Northwestern University McCormick School of Engineering and Applied Science, Chicago, IL, USA

**Keywords:** ventilator-associated pneumonia, mechanical ventilation, machine learning

## Abstract

**Purpose:** Ventilator-associated pneumonia (VAP) remains one of the most serious hospital-acquired infections in the intensive care unit (ICU), with high morbidity and mortality. Early identification of patients at risk for developing VAP could enable timely diagnostics and intervention. However, current clinical tools are limited in their ability to detect early physiologic signals preceding VAP onset. We aimed to build supervised machine learning models to predict short term onset of VAP.

**Methods:** We analyzed electronic health record data from a prospective observational cohort of ICU patients, where VAP was adjudicated using a standardized published protocol by a panel of critical care physicians. Clinical features (including vital signs, ventilator settings, laboratory values, and support devices) were extracted for each patient-ICU-day. We explored unsupervised clustering to characterize feature dynamics associated with VAP onset. We built multiple machine learning models across different prediction windows (3, 5, 7 days before VAP). We examined model performance in two external cohorts, MIMIC-IV and secondary analysis of the AMIKINHAL trial. Results were evaluated with discrimination metrics such as AUROC.

**Results:** The internal cohort included 507 patients with BAL-confirmed diagnoses: 261 developed VAP and 246 did not have VAP. Visualization using clustering identified distinct physiologic states enriched for VAP-labeled days. The best-performing model achieved an AUROC of 0.866 in predicting VAP up to seven days before clinical diagnosis. Temporal model probability trajectories showed rising model confidence in the days leading up to VAP. On external validation in MIMIC-IV, the best model achieved an AUROC of 0.817 for forecasting VAP within five days. There was low feature overlap with the AMIKINHAL trial data, leading to poor model performance. Feature analysis revealed that platelet count, positive end-expiratory pressure (PEEP), ventilator duration, and inflammatory markers were key drivers of model predictions.

**Conclusions:** Machine learning models trained on routinely collected ICU data with careful labeling can anticipate VAP onset up to a week in advance with strong predictive performance. Model performance generalized to data from an entirely different hospital system despite differences in practice and labeling patterns, but did not perform well when there was poor feature overlap. Future work should focus on real-time prospective evaluation.

## Introduction

Ventilator-associated pneumonia (VAP) is a common and serious complication among patients requiring mechanical ventilation [1]. It is associated with increased morbidity, extended intensive care unit (ICU) stays, and elevated mortality risk [2]. Despite widespread implementation of clinical guidelines and prevention strategies, VAP remains difficult to detect early, with diagnosis often delayed until after deterioration has already occurred. Delays can lead to late initiation of therapy and potentially worse outcomes. While some patients show clear clinical signs of infection, others present with more subtle changes in physiology. These changes are often masked by the complex and dynamic nature of critical illness, complicating early recognition of VAP. Current clinical tools lack the temporal use and predictive specificity needed to surface early VAP risk in real time [3]. As a result, clinicians frequently rely on subjective assessments or reactive thresholds, which may not capture emerging infections [4].

Patients in the ICU who require mechanical ventilation are monitored closely for signs of infection, and antibiotics are typically administered if clinical suspicion of pneumonia arises. However, these therapies are administered after clinical deterioration and associated organ dysfunction. Prophylactic and preventive therapies such as inhaled antibiotics [5], selective gastric decontamination [6], and systemic antibiotics [7] to reduce VAP in certain patient populations have been found beneficial in some settings [8]. Widespread use of these therapies may result in antibiotic complications and the emergence of antimicrobial pathogens. A risk stratification strategy for VAP using electronic health record (EHR) predictive models could identify high risk patients and assist clinicians in the judicious use of prophylactic therapies to prevent VAP.

Recent advances in machine learning (ML) have shown promise in critical care, including prediction of sepsis [9], respiratory failure [10], and mortality [11]. However, few studies have focused specifically on early VAP prediction. Moreover, existing models use static features or coarse temporal resolutions [12], limiting their clinical utility. We hypothesized that using high quality ICU data including gold-standard VAP diagnosis based on bronchoalveolar lavage (BAL) fluid sampling and clinical adjudication, we could develop machine learning models to forecast VAP sufficiently early in the patient’s ICU course to be clinically actionable. Some of this work was previously presented at the 2025 American Thoracic Society Meeting [13].

## Methods

### Cohort and Data Source

We analyzed data from patients from the Successful Clinical Response in Pneumonia Therapy (SCRIPT) cohort [14], a prospective observational study of patients on mechanical ventilation who undergo a BAL for suspected pneumonia. We included data from June 2018 up to July 2023. Patients’ families or authorized representatives consent to participate in the study, which is approved by the Northwestern IRB (STU00204868). We used the *CarpeDiem* dataset [15], a structured, day-level ICU database that includes vitals, laboratory measurements, support measures, medications, BAL results, and outcome annotations for SCRIPT patients. Data were abstracted from the electronic health record and study REDCap forms as previously described [16].

### Pneumonia Adjudication and Labels

At our hospital, clinicians routinely obtain BAL to evaluate for pneumonia early on in clinical decompensation. Thus, we annotate the start of a pneumonia episode to correspond with the day of the BAL. BAL results were used to define bacterial VAP, based on microbiology results and the presence of alveolar neutrophilia, along with clinical systemic features such as leukocytosis, fever, and new chest imaging findings. Additional details of adjudication using a standardized multi-reviewer adjudication process are described in our previous work [17]. To model the onset of VAP, using the clearest cases, we developed labels based on the temporal relationship between intubated days and BAL-confirmed events (Figure 1A). As VAP can only develop in intubated patients, we only used intubated days for modeling. Intubated days occurring 3, 5, or 7 days prior to a confirmed VAP or non-VAP event were labeled as positive/negative, respectively. Additional details on labeling are available in the Supplement.

**Figure 1:**
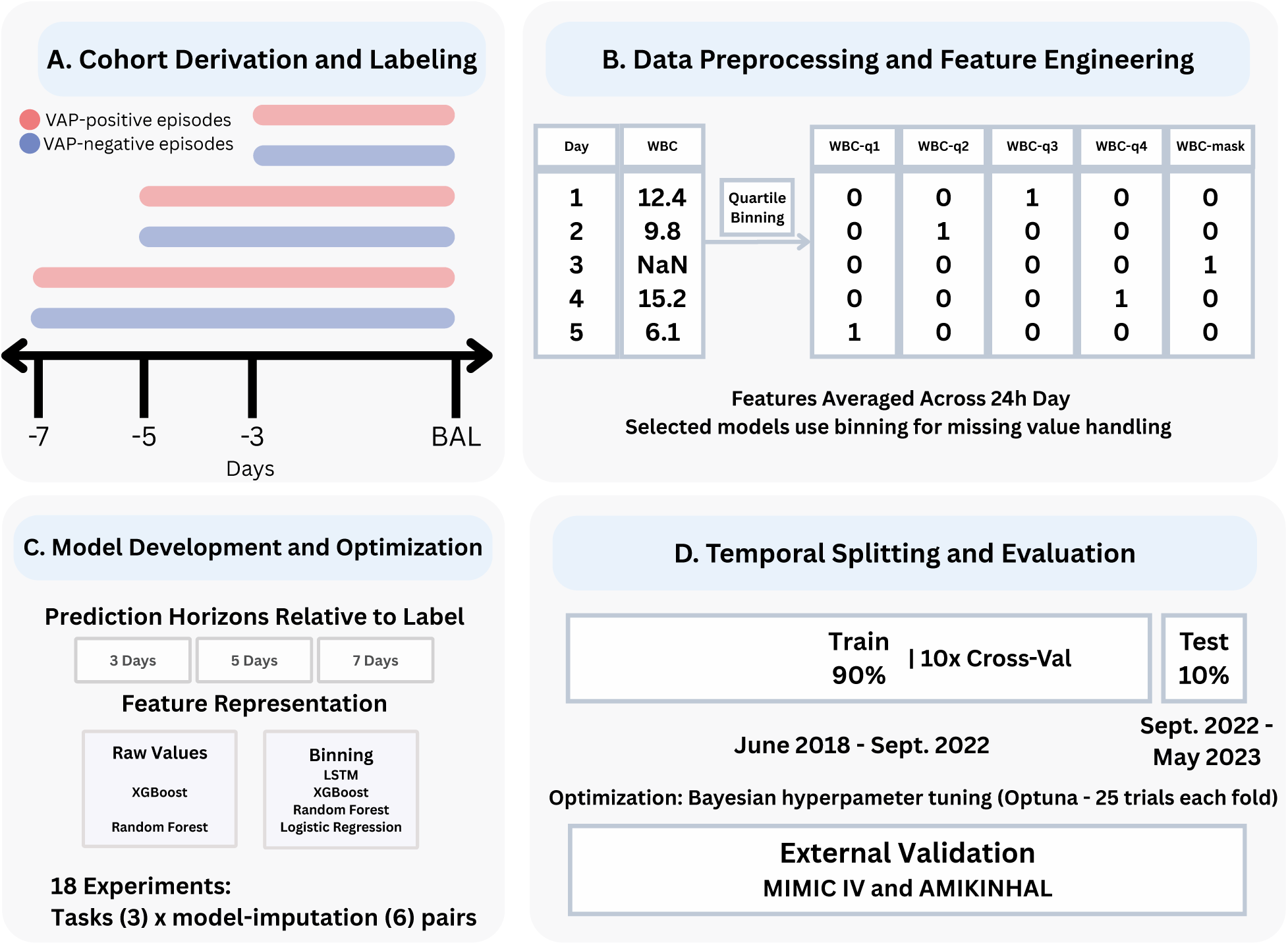
Overview of the machine learning pipeline for ventilator-associated pneumonia (VAP) onset prediction. (A) Cohort derivation and labeling from BAL-confirmed VAP or non-VAP cases with 3-, 5-, and 7-day windows. (B) Data preprocessing with feature extraction, normalization, and missingness handling. (C) Model development using logistic regression, random forest, and XGBoost with Optuna tuning. (D) Temporal splitting and evaluation: 10-fold patient-grouped cross-validation on training set with hyperparameter optimization. Temporal train/test split (90/10; test = most recent patients). Within training, 10-fold GroupKFold CV prevents patient leakage. Each fold optimizes hyperparameters (25 Optuna trials) and evaluates on fold validation data. The hyperparameter set achieving the highest mean validation AUC across all folds is selected, and a final model is trained on all training data using these selected hyperparameters and evaluated on the temporal test set.

### Features

Our model utilized 36 clinical features spanning demographics, vital signs, neurological status, respiratory mechanics, blood gas values, hematology, blood chemistry, inflammatory markers, medications and interventions, and critical care flags. A full list of features is provided in the Supplement. Multiple measurements from the same day were averaged. We performed basic outlier checks, unit harmonization, and quality checks as previously described [16].

### Missing Data

Data missingness is a characteristic of real-world EHR data [18]. Importantly, these missing data do not occur at random, and in many cases, reflect a change in a patient’s clinical status that influences how often a clinician decides to monitor their values or are inherent to their clinical status (i.e. an absence of ventilator features once the patient has been extubated, an improved patient stops having lactate examined). The proportion of missingness across features are summarized in Supplemental Table S1. The models we used are either inherently capable of handling missing data such as XGBoost, or we performed quartile binning for continuous variables, mask encoding for missingness, and one-hot encoding for categorical variables (Figure 1B) [19, 20].

### SOFA trends to predict VAP

Previous work suggested that trends in Sequential Organ Failure Assessment (SOFA) scores could predict the development of subsequent VAP [21]. We examined this by calculating the change in SOFA score between the first and second intubated days for each patient and examining its relationship with subsequent VAP.

### Unsupervised Phenotyping and Temporal Dynamics

We explored latent clinical states associated with or without subsequent VAP through unsupervised clustering of daily patient-level feature vectors. Each data point represented a single intubated day and 36 features. Hierarchical clustering with Ward’s linkage was used to generate patient-day clusters, as described in previous work [16]. Dimensionality reduction with Uniform Manifold Approximation and Projection (UMAP) was applied to visualize cluster structure and the distribution of VAP and non-VAP days in the embedded space.

### Temporal Feature Dynamics

To characterize deterioration in the days preceding VAP onset, we identified patients with exactly five consecutive intubated days leading up to their first BAL-confirmed pneumonia event. The 36 clinical features were extracted and z-score normalized across the cohort. We computed the average value of each feature by day (from Day *−*5 to Day *−*1) and visualized this using a heatmap of feature intensities over time. In addition to magnitude-based trends, we calculated slopes to analyze the steepness of change for each feature between consecutive days.

### Machine Learning, Model Optimization, Feature Importance

To assess predictive performance, we implemented a machine learning framework (Figure 1C). For each VAP/non-VAP prediction horizon, data were split temporally, with 90% of samples used for model development and 10% held out for testing. Within the training set, logistic regression [22], random forest [23], and XGBoost [24] classifiers were trained using 10-fold patient-level cross-validation, ensuring that no patient appeared in multiple folds. For these models, feature vectors corresponded to single patient-day observations. In parallel, we trained a recurrent neural network based on a long short-term memory (LSTM) [25] architecture to explicitly model longitudinal temporal dependencies. For the LSTM, each patient was represented as a time-ordered sequence of daily feature vectors, and predictions were generated using all available historical observations up to the prediction horizon. Because each patient was assigned a single outcome class (VAP or non-VAP), outcome labels were masked from all model input features to prevent patient-level leakage, particularly in temporal models. Patient-level splits identical to those used for the snapshot-based models were applied to prevent information leakage across sequences. Hyperparameter optimization for all models was performed using Bayesian optimization with Optuna [26], with 50 trials per model per task (Figure 1D). Model performance was evaluated on the held-out test set using standard classification metrics, including area under the receiver operating characteristic curve (AUROC), area under the precision-recall curve (AUPRC), accuracy, F1 score. Ninety-five percent confidence intervals for all metrics were calculated using bootstrap resampling with 1000 iterations [27]. To assess how model confidence evolved over time, we plotted predicted VAP probabilities from Days *−*5 to *−*1 for patients in the test set. Day-wise means and standard deviations were computed and visualized to assess whether model probabilities increased in the days preceding a VAP event. To interpret model predictions, we assessed feature importance using model-specific approaches. For tree-based models (XGBoost), we computed SHapley Additive exPlanations (SHAP) values on held-out data to quantify the contribution of each feature to predicted VAP risk. For other models, feature importance was derived using standard model importance functions, including coefficient magnitudes for logistic regression and permutation-based importance for random forest. All interpretability analyses were performed on held-out test data. We followed the TRIPOD-AI checklist [28], provided in the Supplementary Materials.

### External Validation

To examine the performance of our models in external datasets, we used data from MIMIC-IV [29], which includes ICU admissions from the Beth Israel Deaconess Medical Center from 2008-2019. We labeled VAP cases using similar logic as Agard et al. [30], where suspected infection is flagged as the start of antibiotics and obtaining of a respiratory specimen, in patients who have been on mechanical ventilation for greater than 48 hours and had an ICD code for pneumonia. We labeled non-VAP cases as those who did not fulfill the suspected infection criteria (antibiotics and respiratory specimen), and did not have an ICD code for pneumonia; despite having been on mechanical ventilation for greater than 48 hours. We also evaluated model performance using data from the AMIKINHAL study, a randomized controlled trial to study the effects of inhaled amikacin on preventing VAP [5]. They used a standardized case report form to collect clinical data and had central adjudication process for VAP, described in their paper in detail. The summary metric also differed at times. Our model used features averaged over the day, whereas in AMIKINHAL variables like temperature were summarized using daily maximum and minimum values, resulting in a data shift. Features unavailable in external datasets were marked as not available in our dataset. Additional details on external datasets and data processing are available in the Supplement.

### Data and Code Availability

A significant portion of the SCRIPT data has already been made available through PhysioNet at https://physionet.org/content/script-carpediem-dataset/1.8.0/ [15]; a future update will include the new patients enrolled since the publication of the original dataset. The MIMIC-IV dataset is available through PhysioNet to credentialed users who sign a DUA at https://physionet.org/content/mimiciv/3.1/ [29]. The AMIKINHAL dataset was made available to us by the authors after signing a DUA [5]. Code to reproduce our analysis is available at https://github.com/NUSCRIPT/VAPOnset.

## Results

### Cohort

The SCRIPT cohort contains a total of 704 mechanically ventilated patients. Of the 704 mechanically ventilated patients in the SCRIPT cohort, 261 experienced a BAL-confirmed episode of VAP, while 246 patients underwent BAL for suspected pneumonia but were adjudicated to not have VAP (‘non-VAP’). Detailed inclusion criteria, episode adjudication, and exclusion numbers are described in the Supplement. Pathogen results from VAPs are presented in Supplemental Table S2. A flow diagram of cohort inclusion and labeling is provided in Figure S1, and example pneumonia episode timelines are shown in Supplemental Figure S2.

Baseline demographic and clinical outcomes of the VAP and non-VAP cohorts are presented in Table 1. Compared to the non-VAP group, VAP patients had significantly longer ICU stays (median 28.0 vs. 9.0 days, *p <* 0.001), greater ventilator duration (median 27.0 vs. 6.0 days, *p <* 0.001), and increased rates of ECMO use (33.7% vs. 11.8%, *p <* 0.001) and tracheostomy (51.0% vs. 9.3%, *p <* 0.001). Discharge to home was less common in the VAP group (13.4% vs. 32.9%, *p <* 0.001).

**Table 1:** Baseline demographics and clinical characteristics stratified by Ventilator-Associated Pneumonia (VAP) status. P-values were calculated using Mann–Whitney U tests for non-normally distributed continuous variables and chi-square tests for categorical variables. Abbreviations: BMI, body mass index; SOFA, Sequential Organ Failure Assessment; NAT, net antimicrobial therapy; CRRT, continuous renal replacement therapy; ECMO, extracorporeal membrane oxygenation; ICU, intensive care unit; LTACH, long-term acute care hospital; SNF, skilled nursing facility.

| Characteristic | Overall | Did not have VAP | Had VAP | P-Value |
| --- | --- | --- | --- | --- |
| n | 507 | 246 | 261 |  |
| Age (years), median [Q1,Q3] | 62.0 [51.0,71.0] | 61.5 [50.0,72.0] | 62.0 [51.0,70.0] | 0.583 |
| Gender, n (%) |  |  |  |  |
| Female | 189 (37.3) | 110 (44.7) | 79 (30.3) | <0.001 |
| Male | 318 (62.7) | 136 (55.3) | 182 (69.7) |  |
| Race, n (%) |  |  |  |  |
| White | 310 (61.1) | 159 (64.6) | 151 (57.9) | 0.292 |
| Black or African American | 103 (20.3) | 49 (19.9) | 54 (20.7) |  |
| Asian | 15 (3.0) | 7 (2.8) | 8 (3.1) |  |
| Unknown/Not Reported | 79 (15.6) | 31 (12.6) | 48 (18.4) |  |
| Ethnicity, n (%) |  |  |  |  |
| Hispanic or Latino | 94 (18.5) | 31 (12.6) | 63 (24.1) | 0.003 |
| Not Hispanic or Latino | 389 (76.7) | 204 (82.9) | 185 (70.9) |  |
| Unknown/Not Reported | 24 (4.7) | 11 (4.5) | 13 (5.0) |  |
| BMI, median [Q1,Q3] | 28.2 [24.2,33.4] | 27.3 [23.3,33.0] | 29.1 [25.0,34.0] | 0.060 |
| Smoking Status, n (%) |  |  |  |  |
| Never Smoked | 219 (43.2) | 108 (43.9) | 111 (42.5) | 0.066 |
| Former Smoker | 126 (24.9) | 70 (28.5) | 56 (21.5) |  |
| Current Smoker | 41 (8.1) | 21 (8.5) | 20 (7.7) |  |
| Unknown | 121 (23.9) | 47 (19.1) | 74 (28.4) |  |
| Immunocompromised, n (%) | 118 (23.3) | 61 (24.8) | 57 (21.8) | 0.495 |
| Admission SOFA Score, median [Q1,Q3] | 5.0 [2.0,10.0] | 5.0 [2.0,10.0] | 6.0 [2.0,11.0] | 0.074 |
| Cumulative NAT Score, median [Q1,Q3] | 0.0 [-17.0,16.5] | -3.0 [-18.0,7.0] | 5.0 [-14.0,25.0] | <0.001 |
| Cumulative Steroid Dose, median [Q1,Q3] | 600.0 [0.0,1840.0] | 550.0 [0.0,1545.0] | 640.0 [0.0,2050.0] | 0.292 |
| Norepinephrine use, n (%) | 438 (86.4) | 194 (78.9) | 244 (93.5) | <0.001 |
| Hemodialysis, n (%) | 66 (13.0) | 20 (8.1) | 46 (17.6) | 0.002 |
| CRRT, n (%) | 145 (28.6) | 54 (22.0) | 91 (34.9) | 0.002 |
| ECMO use, n (%) | 117 (23.1) | 29 (11.8) | 88 (33.7) | <0.001 |
| Total ICU Days, median [Q1,Q3] | 17.0 [8.0,31.5] | 9.0 [6.0,17.8] | 28.0 [16.0,44.0] | <0.001 |
| Tracheostomy, n (%) | 156 (30.8) | 23 (9.3) | 133 (51.0) | <0.001 |
| Total Intubation Days, median [Q1,Q3] | 12.0 [5.0,28.0] | 6.0 [3.0,12.0] | 27.0 [13.0,39.0] | <0.001 |
| Discharge Disposition, n (%) |  |  |  |  |
| Home | 116 (22.9) | 81 (32.9) | 35 (13.4) | <0.001 |
| Rehab | 95 (18.7) | 57 (23.2) | 38 (14.6) |  |
| LTACH | 72 (14.2) | 16 (6.5) | 56 (21.5) |  |
| SNF | 33 (6.5) | 22 (8.9) | 11 (4.2) |  |
| Hospice | 16 (3.2) | 15 (6.1) | 1 (0.4) |  |
| Died | 175 (34.5) | 55 (22.4) | 120 (46.0) |  |
| Unfavorable Outcome, n (%) | 191 (37.7) | 70 (28.5) | 121 (46.4) | <0.001 |

### Unsupervised Clustering Analysis

Using a six-cluster strategy, we observed that VAP-labeled days were highly enriched within specific clusters characterized by elevated inflammatory markers (e.g., C-reactive protein, ferritin, neutrophil count), impaired gas exchange (e.g., elevated PaCO_2_), and increased ventilatory support requirements (Figure 2A).

**Figure 2:**
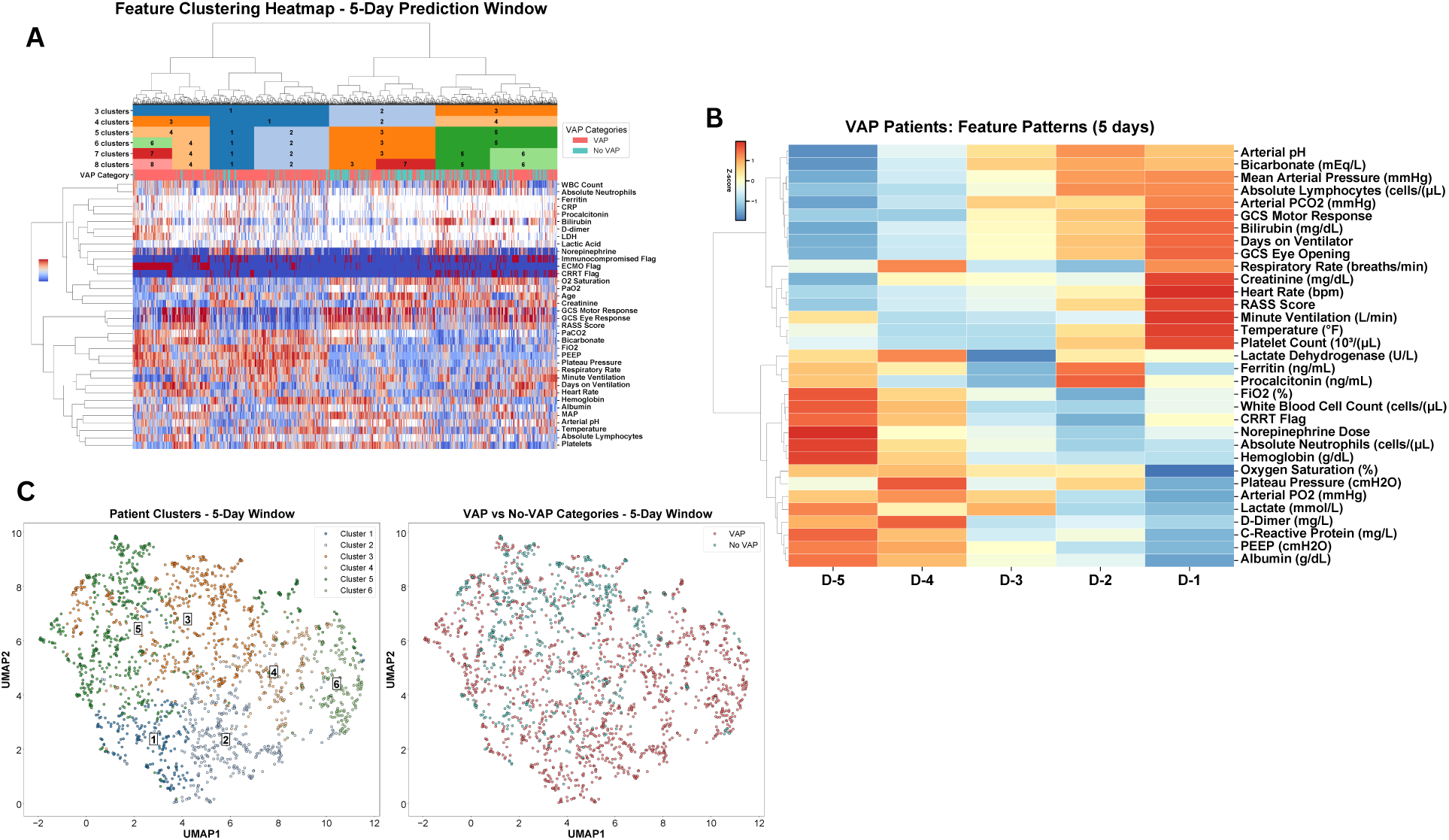
Unsupervised clustering reveals physiologic states associated with impending VAP. (**A**) Similarity-based clustering of patient-days over a 5-day prediction window identifies distinct physiologic clusters, with VAP-labeled days enriched in clusters marked by heightened inflammation and increased ventilatory support. (**B**) Aggregated temporal trajectories of key clinical variables in the five days preceding BAL-confirmed VAP demonstrate coordinated shifts across inflammatory, hemodynamic, and metabolic features. (**C**) UMAP projection of patient-day feature vectors shows spatial concentration of VAP days within specific regions of the embedded space, despite pneumonia labels not being used during clustering.

To further assess global structure, patient-day representations were projected into two dimensions using Uniform Manifold Approximation and Projection (UMAP). Despite pneumonia labels not being used during clustering, VAP days localized to distinct regions of the UMAP, indicating coherent physiologic states associated with infection (Figure 2C).

To characterize temporal evolution preceding VAP diagnosis, we analyzed a subset of 85 patients with seven consecutive intubated days prior to their first BAL-confirmed infection. Aggregated trajectories across the cohort revealed systematic shifts in multiple nonrespiratory parameters, including increases in bilirubin, heart rate, temperature, absolute lymphocyte count, and mean arterial pressure from Day *−*5 to Day *−*1, alongside declines in CRRT use, procalcitonin, D-dimer, and lactate dehydrogenase (Figure 2B).

### Early SOFA Change

The distribution of SOFA score changes was similar across VAP and non-VAP groups, and no statistically significant difference was observed. SOFA difference yielded a low AUC of 0.528 at predicting VAP development in our cohort (Supplemental Figure S3).

### Supervised Machine Learning Performance

We implemented a full modeling pipeline using ten-fold patient-level cross-validation and Optuna-based hyperparameter tuning. LSTM models achieved AUCs up to 0.866 for 7 day prediction tasks and average precision scores reaching 0.903 and Random Forest with raw imputation achieved 0.843 AUC with 5 day prediction tasks (Table 2). Model calibration assessed by plotting observed versus predicted VAP risk and summarized using Brier scores (0.1518-0.1721) is provided in Supplemental Figure S4. It is important to note that because prediction datasets differed across the 3-, 5-, and 7-day horizons, AUROC and AUPRC values across horizons are not directly comparable as they were evaluated on non-identical test sets.

**Table 2:** Internal testing performance with 95% confidence intervals. Baseline AUPRCs were 0.695 (3-day), 0.683 (5-day), and 0.618 (7-day), reflecting outcome prevalence in the training dataset.

| Task Model | AUC | AUPRC | Accuracy | F1 |
| --- | --- | --- | --- | --- |
| *VAP Within 3 Days* |  |  |  |  |
| XGBoost (raw) | 0.721 (0.626–0.797) | 0.878 (0.819–0.921) | 0.625 (0.539–0.703) | 0.711 (0.622–0.779) |
| **Random Forest (raw)** | **0.806 (0.732–0.874)** | **0.918 (0.876–0.952)** | **0.750 (0.672–0.820)** | **0.828 (0.766–0.881)** |
| XGBoost (binned) | 0.756 (0.670–0.830) | 0.899 (0.849–0.937) | 0.656 (0.578–0.734) | 0.711 (0.619–0.784) |
| Random Forest (binned) | 0.769 (0.682–0.844) | 0.902 (0.854–0.941) | 0.695 (0.609–0.773) | 0.775 (0.699–0.835) |
| Logistic Regression (binned) | 0.725 (0.637–0.809) | 0.881 (0.826–0.927) | 0.664 (0.578–0.742) | 0.749 (0.671–0.815) |
| LSTM (binned) | 0.765 (0.676–0.850) | 0.899 (0.844–0.943) | 0.617 (0.531–0.703) | 0.642 (0.543–0.731) |
| *VAP Within 5 Days* |  |  |  |  |
| XGBoost (raw) | 0.706 (0.634–0.772) | 0.871 (0.826–0.911) | 0.613 (0.543–0.678) | 0.680 (0.612–0.744) |
| **Random Forest (raw)** | **0.843 (0.782–0.894)** | **0.931 (0.896–0.959)** | **0.789 (0.729–0.844)** | **0.842 (0.789–0.887)** |
| XGBoost (binned) | 0.747 (0.677–0.807) | 0.891 (0.851–0.926) | 0.658 (0.588–0.719) | 0.728 (0.661–0.787) |
| Random Forest (binned) | 0.780 (0.715–0.838) | 0.905 (0.866–0.938) | 0.729 (0.663–0.784) | 0.799 (0.740–0.846) |
| Logistic Regression (binned) | 0.716 (0.643–0.781) | 0.869 (0.823–0.911) | 0.653 (0.588–0.714) | 0.732 (0.669–0.786) |
| LSTM (binned) | 0.840 (0.782–0.892) | 0.928 (0.893–0.957) | 0.774 (0.714–0.829) | 0.812 (0.756–0.861) |
| *VAP Within 7 Days* |  |  |  |  |
| XGBoost (raw) | 0.812 (0.757–0.863) | 0.904 (0.867–0.933) | 0.745 (0.687–0.795) | 0.766 (0.703–0.818) |
| Random Forest (raw) | 0.846 (0.799–0.889) | **0.916 (0.880–0.943)** | 0.764 (0.714–0.811) | **0.817 (0.769–0.858)** |
| XGBoost (binned) | 0.816 (0.760–0.866) | 0.906 (0.869–0.936) | 0.761 (0.707–0.811) | 0.780 (0.720–0.829) |
| Random Forest (binned) | 0.826 (0.773–0.876) | 0.910 (0.874–0.938) | 0.737 (0.680–0.788) | 0.793 (0.738–0.839) |
| Logistic Regression (binned) | 0.792 (0.735–0.843) | 0.890 (0.850–0.923) | 0.687 (0.629–0.741) | 0.746 (0.690–0.798) |
| **LSTM (binned)** | **0.866 (0.820–0.909)** | 0.904 (0.846–0.948) | **0.772 (0.722–0.822)** | 0.803 (0.747–0.852) |

### Feature Importance and Model Interpretability

Feature rankings were largely consistent across prediction horizons and modeling approaches, with ventilator mechanics, inflammatory markers, and oxygenation-related variables repeatedly emerging as key contributors to VAP risk (Figure 3A-B). Measures reflecting ventilator exposure and respiratory support, including days on ventilator, plateau pressure, positive end-expiratory pressure (PEEP), and fraction of inspired oxygen (FiO_2_), were among the most influential features across models. Additional contributions from arterial blood gas parameters, platelet count, and inflammatory markers suggest that evolving physiologic and systemic changes precede clinical diagnosis of VAP.

**Figure 3:**
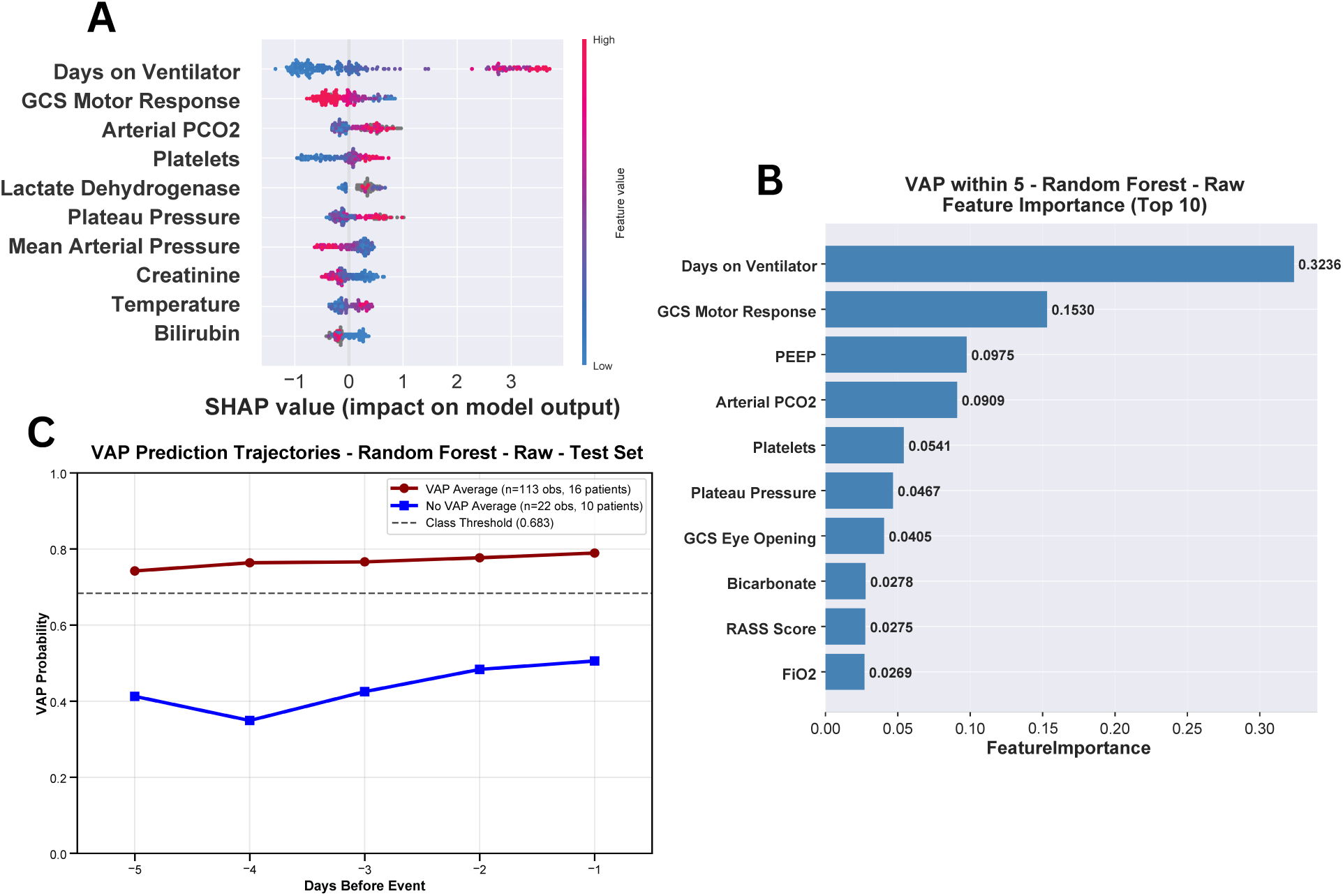
Model interpretability and temporal risk dynamics for VAP prediction. (**A**) SHAP summary plot for the XGBoost model predicting VAP within five days, illustrating the direction and magnitude of feature contributions across patient-days. Higher values of ventilatory support variables (e.g., days on ventilator, plateau pressure) and impaired neurologic status were associated with increased predicted VAP risk. (**B**) Feature importance rankings from a Random Forest model trained on the same prediction task, demonstrating similar prioritization. (**C**) Temporal evolution of predicted VAP risk probabilities from Day *−*5 to Day *−*1 prior to diagnosis.

### Temporal Rise in Predicted VAP Risk

To assess whether model predictions tracked temporal progression toward infection, we examined predicted probabilities for patients with exactly one intubated observation per day from Day *−*5 to Day *−*1 relative to VAP onset. Among patients who subsequently developed VAP, predicted risk increased steadily over time, rising from a mean probability of approximately 75.0% on Day *−*5 to nearly 79.0% by Day *−*1 (Figure 3C). In contrast, predicted probabilities for non-VAP patients remained consistently low and showed minimal temporal variation over the same period.

### External Validation on the MIMIC-IV Cohort and AMIKINHAL trial

External validation was performed using the MIMIC-IV cohort, which differed substantially from the internal SCRIPT cohort. The MIMIC-IV cohort was slightly older, predominantly male, and presented a much lower prevalence of VAP alongside substantially higher overall mortality, reflecting differences ICU exposure and competing risks between cohorts (Table S3, Table S4).(Supplemental Figure S5, Table S3, Table S4). The MIMIC-IV cohort contained a markedly higher proportion of non-VAP cases, resulting in low baseline AUPRCs of 0.075, 0.104, and 0.123 for the 3-, 5-, and 7-day prediction windows, respectively (Table 3). Despite these differences, models trained on raw continuous features demonstrated stable discriminative performance across prediction horizons, with AUC values reaching *>* 0.80 for all tasks. In contrast, models trained on binned features showed reduced performance in terms of AUPRC and F1 score, particularly for longer prediction windows. Binning thresholds were derived from the internal SCRIPT cohort and applied directly to the external MIMIC-IV data, where shifted feature distributions led to compression of many values into a limited number of bins (Supplemental Figure S6).

**Table 3:** External validation performance on the MIMIC-IV cohort with 95% confidence intervals. Baseline AUPRCs were 0.0675 (3-day), 0.0944 (5-day), and 0.1126 (7-day), reflecting outcome prevalence in the external dataset.

| Task Model | AUC | AUPRC | Accuracy | F1 |
| --- | --- | --- | --- | --- |
| *VAP Within 3 Days (External)* |  |  |  |  |
| XGBoost (raw) | **0.807 (0.783–0.829)** | 0.347 (0.305–0.396) | 0.341 (0.329–0.353) | 0.172 (0.158–0.187) |
| Random Forest (raw) | 0.800 (0.776–0.822) | **0.365 (0.320–0.411)** | 0.112 (0.105–0.119) | 0.144 (0.132–0.155) |
| XGBoost (binned) | 0.711 (0.684–0.737) | 0.315 (0.270–0.360) | 0.826 (0.817–0.836) | 0.275 (0.243–0.304) |
| Random Forest (binned) | 0.664 (0.636–0.692) | 0.262 (0.223–0.305) | 0.500 (0.487–0.513) | 0.173 (0.156–0.189) |
| Logistic Regression (binned) | 0.658 (0.631–0.684) | 0.206 (0.172–0.244) | 0.601 (0.589–0.613) | 0.190 (0.171–0.208) |
| LSTM | 0.652 (0.621–0.681) | 0.225 (0.188–0.269) | **0.863 (0.855–0.871)** | **0.296 (0.262–0.331)** |
| *VAP Within 5 Days (External)* |  |  |  |  |
| XGBoost (raw) | 0.803 (0.785–0.823) | 0.449 (0.413–0.489) | 0.412 (0.400–0.424) | 0.241 (0.225–0.256) |
| Random Forest (raw) | **0.817 (0.796–0.838)** | **0.486 (0.447–0.530)** | 0.209 (0.200–0.219) | 0.202 (0.190–0.215) |
| XGBoost (binned) | 0.658 (0.635–0.682) | 0.289 (0.253–0.327) | 0.707 (0.695–0.718) | 0.262 (0.240–0.286) |
| Random Forest (binned) | 0.658 (0.634–0.683) | 0.352 (0.314–0.389) | 0.558 (0.545–0.571) | 0.224 (0.207–0.244) |
| Logistic Regression (binned) | 0.627 (0.607–0.649) | 0.145 (0.131–0.161) | 0.564 (0.552–0.576) | 0.230 (0.211–0.248) |
| LSTM | 0.662 (0.634–0.687) | 0.353 (0.312–0.391) | **0.846 (0.838–0.855)** | **0.354 (0.321–0.383)** |
| *VAP Within 7 Days (External)* |  |  |  |  |
| XGBoost (raw) | **0.813 (0.795–0.831)** | **0.559 (0.526–0.595)** | 0.507 (0.495–0.519) | 0.301 (0.283–0.318) |
| Random Forest (raw) | 0.770 (0.753–0.789) | 0.425 (0.392–0.458) | 0.208 (0.199–0.218) | 0.231 (0.217–0.244) |
| XGBoost (binned) | 0.697 (0.676–0.719) | 0.410 (0.374–0.446) | **0.826 (0.818–0.836)** | **0.386 (0.359–0.413)** |
| Random Forest (binned) | 0.705 (0.685–0.725) | 0.405 (0.373–0.439) | 0.674 (0.664–0.685) | 0.314 (0.293–0.334) |
| Logistic Regression (binned) | 0.707 (0.685–0.728) | 0.436 (0.400–0.470) | 0.750 (0.740–0.760) | 0.352 (0.328–0.376) |
| LSTM | 0.637 (0.614–0.660) | 0.337 (0.305–0.370) | 0.769 (0.759–0.779) | 0.318 (0.294–0.342) |

Data in the AMIKINHAL trial had been collected using manual case report forms and had only 9 (out of our 36) features (WBC, CRRT, FiO2, O2 saturation, PaO2, creatinine, PEEP, days on ventilator, temperature). They had additional features that were not captured as structured data in SCRIPT such as tracheal secretions, and radiological infiltrates. We examined feature distributions among these features in comparison with the SCRIPT cohort (Supplemental Figure S7). While several features showed statistically significant differences between VAP and non-VAP groups in both cohorts, effect sizes were substantially smaller in AMIKINHAL. For example, days on ventilator showed a large effect (Cohen’s d = 0.736) in SCRIPT but a negligible effect (d = -0.103) in AMIKINHAL. Using our models to test on the AMIKINHAL data resulted in a 0.6065 [0.5792, 0.6335] AUC for predicting VAP within 3 days.

### Feature Importance in the External MIMIC-IV Cohort

To assess whether the predictors driving model decisions generalized beyond the internal SCRIPT cohort, we examined feature importance for raw-feature models evaluated on the external MIMIC-IV cohort (Supplemental Figure S8). Across both cohorts, duration of mechanical ventilation was the strongest predictor of VAP risk, with neurologic status measures such as GCS motor response, ventilatory mechanics including plateau pressure and PEEP, and laboratory and physiologic variables like arterial PCO_2_, platelet count, bicarbonate, and inflammatory markers also ranking highly.

## Discussion

Despite significant improvements in ICU care and infection control practices, ventilator-associated pneumonia (VAP) remains a frequent and serious complication in mechanically ventilated patients. Although preventive bundles are widely implemented, clinical tools to anticipate VAP onset are lacking. In this study, we developed ML models that use structured, day-level ICU data to predict VAP onset up to seven days in advance. Our approach demonstrates that EHR-based features can be used to detect subtle, evolving clinical signals that precede pneumonia, enabling more proactive risk assessment and decision-making.

We show that unsupervised clustering of patient-days revealed latent phenotypic patterns associated with VAP onset. These clusters were enriched for elevated inflammatory markers, increased ventilator pressures, and impaired gas exchange, consistent with the clinical profile of early pneumonia. Our temporal profiling analysis further supports this, showing that respiratory and inflammatory features begin diverging from baseline up to five days before diagnosis, with progressively increasing trajectories. These changes were not captured by coarse measures like early SOFA score changes alone, which lacked predictive utility, highlighting the need for more granular models in this domain.

Previous efforts to predict VAP using EHR data have been limited by small sample sizes [31], delayed and coarse outcome labels such as ICD codes [12], and models that rely on static snapshots (i.e. using data from the beginning of admission only) [32] rather than dynamic patient trajectories (our day-by-day prediction window). There is also limited external validation in the current studies, with many developed exclusively using the MIMIC-IV dataset [12, 32–34]. Our study addresses these limitations by leveraging a large cohort of intubated patients with state-of-the-art BAL-confirmed and clinically-adjudicated infection status. We used high-resolution temporal data and performed additional unsupervised techniques and feature evaluation to understand physiologic changes over time, and examined our model performance in external datasets. In the nature of open science and rigorous reproducibility, we furthermore fully share our code for evaluation and understanding of detailed decisions made during data processing.

After tuning with Optuna and evaluating multiple modeling strategies across different labeling windows, predictive performance obtained high AUROCs *>* 0.85. SHAP analysis revealed clinically plausible predictors of VAP, including platelet count, PEEP, FiO_2_, and procalcitonin, many of which have mechanistic relevance to pulmonary infection. Interestingly, several features identified as influential to the model, such as platelet count, often still appeared within the normal laboratory-defined ranges and therefore may be overlooked in busy clinical workflows. These findings highlight the value of ML in surfacing early, multivariate signals that may not reach conventional thresholds for clinical concern.

Importantly, our models demonstrated robust performance in an external validation cohort derived from MIMIC-IV, despite substantial differences in outcome prevalence, patient population, and feature distributions. While the prevalence of VAP in MIMIC-IV was considerably lower than in the internal SCRIPT cohort, resulting in lower baseline AUPRC values, raw-feature models maintained strong discriminative ability across prediction horizons. Feature importance patterns in the external MIMIC-IV cohort closely mirrored those observed internally. Duration of mechanical ventilation, neurologic status, and ventilatory mechanics again emerged as the dominant predictors of VAP risk. The strong concordance in feature importance between cohorts supports the generalizability of the learned representations and suggests that the models rely on stable, clinically meaningful signals rather than cohort-specific artifacts. The relatively poor performance of our model in the AMIKINHAL cohort, which lacked many of these important features, supports this interpretation.

This study has several important limitations. Models were built using data from a single center, using a frequent BAL-sampling protocol, contrary to many institutions with different sampling practices, patient populations, or VAP prevention protocols. BAL-confirmed VAP was used as the diagnostic standard along with clinician adjudication; however, clinical variability in when BAL is pursued may introduce selection bias. Our modeling approach excluded patients with unclear adjudication. We used heuristic definitions of VAP/non-VAP in the MIMIC cohort; manual labels by physician experts may have identified different cases of VAP. It is challenging to find external validation cohorts that have both granular EHR data and clean VAP labels. We furthermore relied only on structured data in our models; incorporating unstructured information such as radiology reports or clinical notes via natural language processing may further improve performance. Future work should explore prospective deployment for real-time model evaluation integrated with EHR systems.

## Conclusion

We found that machine learning models can accurately predict the onset of VAP using structured, day-level ICU data routinely captured in clinical practice. By leveraging granular physiologic, laboratory, and ventilator parameters, our models identified subtle but consistent changes in patient status up to seven days prior to clinical diagnosis. In such patients identified by the model to be high-risk for VAP, prophylactic therapies or earlier diagnostics may be beneficial. Prospective validation in diverse, real-time clinical settings will be essential to confirm generalizability and assess implementation impact.

## Supporting information

NU SCRIPT Study Investigators

TRIPOD-AI Checklist

## Data Availability

A significant portion of the SCRIPT data has already been made available through PhysioNet; a future update will include the new patients enrolled since the publication of the original dataset.
The MIMIC-IV dataset is available through PhysioNet to credentialed users who sign a DUA.
The AMIKINHAL dataset was made available to us by the authors after signing a DUA.
Code to reproduce our analysis is available on GitHub.

https://physionet.org/content/script-carpediem-dataset/1.8.0/

https://physionet.org/content/mimiciv/3.1/}

https://www.nejm.org/doi/full/10.1056/NEJMoa2310307

https://github.com/NUSCRIPT/VAPOnset

## Acknowledgements

We would like to thank all the patients and families who consented to take part in the study. Many team members contribute to the SCRIPT Systems Biology Center and are recognized in the NU SCRIPT Study Investigators list. We would like to thank Dr. Geoffray Agard for sharing code to identify suspicion for VAP in the MIMIC-IV cohort. We would like to thank Drs. Anna Barker, Owen Albin, Michael Sjoding, Benjamin Wu, Clea Barnett, and Cecilia Chung and Mr. Mark Nuppnau for looking into potential external datasets in which to validate our model.

## AI use

Claude was used for help coding and ChatGPT was used for help rewording text. The authors reviewed, edited, and take responsibility for all output.

## Supplemental Files

Attached separately:

NU SCRIPT Study Investigators

TRIPOD-AI checklist

## Supplementary Methods

### SCRIPT Cohort Construction, Adjudication, and Episode Filtering

The Successful Clinical Response In Pneumonia Therapy (SCRIPT) cohort has 704 mechanically ventilated patients who underwent bronchoalveolar lavage (BAL) for suspected pneumonia. These patients were recruited from Northwestern Memorial Hospital, a quarternary care hospital in Chicago, IL, USA. Among these, 261 patients were adjudicated as having ventilator-associated pneumonia (VAP) based on BAL-confirmed microbiologic and clinical criteria. Reviewers were not blinded to any clinical results, and had access to the entirety of the patient’s hospital course. An additional 246 patients underwent BAL for suspected pneumonia but were adjudicated post hoc by a multidisciplinary clinical review committee to not represent pneumonia and were designated as non-VAP controls (sepsis from a non-pulmonary source, heart failure, inflammatory process such as ILD flare without evidence of infection, etc). Patients are given a new identifier with each new admission. Patients are followed until the end of each admission. Patient who undergo lung transplant during admission are coded as having died.

In our adjudication process, we label ‘de novo’ VAPs as those occurring after a patient has been off antibiotics for at least 48 hours. ‘Superinfection’ VAPs are those occurring during treatment for a Community-Acquired Pneumonia (HAP) or (Hospital-Acquired Pneumonia) HAP episode. BAL-confirmed pneumonia episodes categorized as community-acquired pneumonia (CAP) or hospital-acquired pneumonia (HAP), in the absence of superinfection or documented microbiologic cure, were identified in 152 patients but excluded from the primary VAP versus non-VAP analyses. CAP/HAP patients who developed ‘superinfection’ (develops while on antibiotic therapy) VAPs were included in analysis as positive VAP cases. CAP/HAP patients that were cured successfully (had resolution of fever, leukocytosis, improvements in ventilator requirements) were included in analysis as non-VAP controls. CAP/HAP patients that had indeterminate cure (adjudicated at day 7, day 10, and day 14) were not included in analysis. Patients with both BAL-confirmed VAP and non-VAP episodes during the same hospitalization (*n* = 15) were excluded to avoid label ambiguity. Additionally, 30 patients who met timing criteria for VAP but whose BAL cultures only recovered only viral pathogens were excluded, as these pathogens were likely not acquired from the hospital/ventilator. Episode duration is linked to duration of treating antibiotics. Other intubated days were excluded from modeling if they occurred within three days of death or lung transplant, overlapped with another VAP episode, or could not be definitively assigned.

A CONSORT-style diagram summarizing cohort construction, exclusions, and final group allocation is provided in Supplemental Figure S1. Timelines illustrating example pneumonia episode types are shown in Supplemental Figure S2. We did not perform any formal sample size calculation; we used all patient data available. Additional of study protocols are available in our previous works [16]; the study was not formally registered. While VAP is known to have worse inequalities in low/middle-income countries, we were not able to subset analyses by this feature due to recruitment and validation cohorts in high income countries [2]. Parts of the SCRIPT protocol were reviewed with the hospital Patient and Family Advisory Committee during study development.

The full list of features included demographics and timeline information such as age at hospital admission and days on ventilator; vital signs including temperature, heart rate, mean arterial pressure, respiratory rate, and oxygen saturation; neurological measures comprising Glasgow Coma Scale eye opening, Glasgow Coma Scale motor response, and RASS score; ventilator parameters such as PEEP, fraction of inspired oxygen (FiO_2_), plateau pressure, and minute ventilation; blood gas values including arterial pH, arterial PCO_2_, and arterial PO_2_; hematologic measures including white blood cell count, absolute lymphocytes, absolute neutrophils, hemoglobin, and platelet count; chemistry variables including bicarbonate, creatinine, albumin, and bilirubin; inflammatory markers including C-reactive protein, D-dimer, ferritin, lactate dehydrogenase, lactic acid, and pro-calcitonin; medications and interventions such as norepinephrine use; and clinical flags indicating immunocompromised status, ECMO use, and CRRT use.

Within the 3-day prediction window, 904 patient-days were labeled as VAP and 337 as non-VAP; within the 5-day window, 1,404 patient-days were labeled as VAP and 565 as non-VAP; and within the 7-day window, 1,815 patient-days were labeled as VAP and 858 as non-VAP. Patients were followed until the end of their hospitalization, with pneumonia episode adjudication for the 99 days post study enrollment.

### External Validation Cohort Construction and Patient Filtering

The first external validation cohort was derived from the MIMIC-IV database, which contains EHR data from ICU admissions at Beth Israel Deaconess Medical Center between 2008 and 2019. Adult ICU admissions were initially identified and subsequently filtered to define a mechanically ventilated cohort at risk for VAP.

Patients were first restricted to those receiving invasive mechanical ventilation for at least 48 hours continuously, consistent with standard definitions of VAP and prior work [30]. ICU admissions not meeting this ventilation duration requirement were excluded.

Eligible ICU encounters were then expanded into a daily patient-day format to support temporal labeling and modeling. Encounters with invalid or incomplete timestamp information preventing alignment of clinical events to ICU days were excluded at this stage.

To identify candidate VAP events, respiratory culture and pneumonia-related events were merged into the daily cohort. Events with invalid timestamps, absent pneumonia labels, or without a valid mapping to an ICU encounter block were excluded. Candidate events were further filtered using strict temporal criteria: respiratory specimens were required to be obtained at least 48 hours after initiation of mechanical ventilation and no more than 48 hours following extubation. Events failing to meet these timing constraints were not considered eligible VAP events.

Daily VAP labels were assigned only on patient-days meeting all eligibility criteria; patient-days failing to meet these criteria were assigned missing values. To ensure a single label per encounter-day, duplicate candidate events occurring on the same encounter-day were resolved by retaining the event with the positive VAP label.

For patient-level classification, individuals were categorized using a first-label approach. Specifically, each patient was assigned to a group based on the earliest VAP label observed during their ICU stay. Patients whose first eligible labeled day corresponded to a VAP were classified as VAP cases, while those whose first eligible labeled day corresponded to a non-VAP were classified as controls. Patients who never received an eligible labeled day throughout their ICU stay were excluded from supervised analyses.

A CONSORT diagram summarizing cohort construction, filtering steps, and final group allocation is provided in Figure S5.

### Features Unavailable in the External MIMIC-IV Cohort

While the majority of 36 clinical features used for model development in the SCRIPT cohort were also available in the MIMIC-IV dataset, a subset of variables could not be derived from the external data and were therefore excluded from external analyses. Specifically, three features present in SCRIPT were unavailable or inconsistently recorded in MIMIC-IV: D-dimer, procalcitonin, and immunocompromised status.

### AMIKINHAL External Validation Cohort Construction and Patient Filtering

The second external validation cohort was derived from the AMIKINHAL clinical trial, a randomized controlled trial evaluating inhaled amikacin for VAP prevention in mechanically ventilated ICU patients. Patient enrollment and data collection details are outlined in their paper.[5] The dataset included 847 patients enrolled in the trial. Daily measurements were recorded in a wide format for each patient over a 28-day period following intubation. VAP was adjudicated as a binary outcome (yes/no) by clinical investigators from day 4 through day 28 of the study period. For modeling purposes, the dataset was converted to a patient-day format, with each row representing a patient on a given day of the 28-day period. The following variables were available: temperature, WBC, PEEP, CRRT flag, PaO2, FiO2, creatinine, SpO2, days on ventilator, and adjudicated VAP outcome. Unit conversions were performed to standardize the dataset: creatinine was converted from µmol/L to mg/dL (conversion factor: 1/88.42), and temperature was converted from Celsius to Fahrenheit. Patient-days were excluded if they occurred after a VAP event, if extubation occurred on that day, or if critical temporal variables (days on ventilator, study day) were missing. For predictive modeling, labels were created using windows of 3, 5, and 7 days preceding VAP events, consistent with the MIMIC cohort approach. Patient-days falling within these windows before an adjudicated VAP event were labeled as 1 (VAP), while patient-days preceding a confirmed non-VAP outcome were labeled as 0 (non-VAP). Patient-days occurring after a VAP event or outside any label window were excluded from analysis.

### Features Unavailable in the External AMIKINHAL Cohort

The AMIKINHAL trial dataset contained only 9 features available in the SCRIPT cohort. All other features from the full feature set were unavailable. Missing vital signs included heart rate, mean arterial pressure, and respiratory rate. Neurological assessments were unavailable, including GCS eye opening, GCS motor response, and RASS scores. Ventilator parameters missing included plateau pressure and minute ventilation. Blood gas values unavailable included arterial pH and PaCO2. Hematologic measures missing included absolute lymphocyte count, absolute neutrophil count, hemoglobin, and platelet count. Chemistry variables unavailable included bicarbonate, albumin, and bilirubin. Inflammatory markers missing included C-reactive protein, D-dimer, ferritin, lactate dehydrogenase, lactic acid, and procalcitonin. Medication records were limited to a binary vasopressor flag rather than specific norepinephrine dosing. Clinical flags unavailable included immunocompromised status and ECMO use. Missing features during testing were denoted as NaN.

### Next steps in model development

The current model requires additional validation with real-time prospective deployment in a system integrated with the EHR. After that, future details on access (such as via API, object, etc) can be considered. Given our models inherently handle missing data, no specific handling of unavailable input data will be performed. We envision limited expertise needed from end-users.

### Supplemental Figures and Tables

**Supplementary Figure S1:**
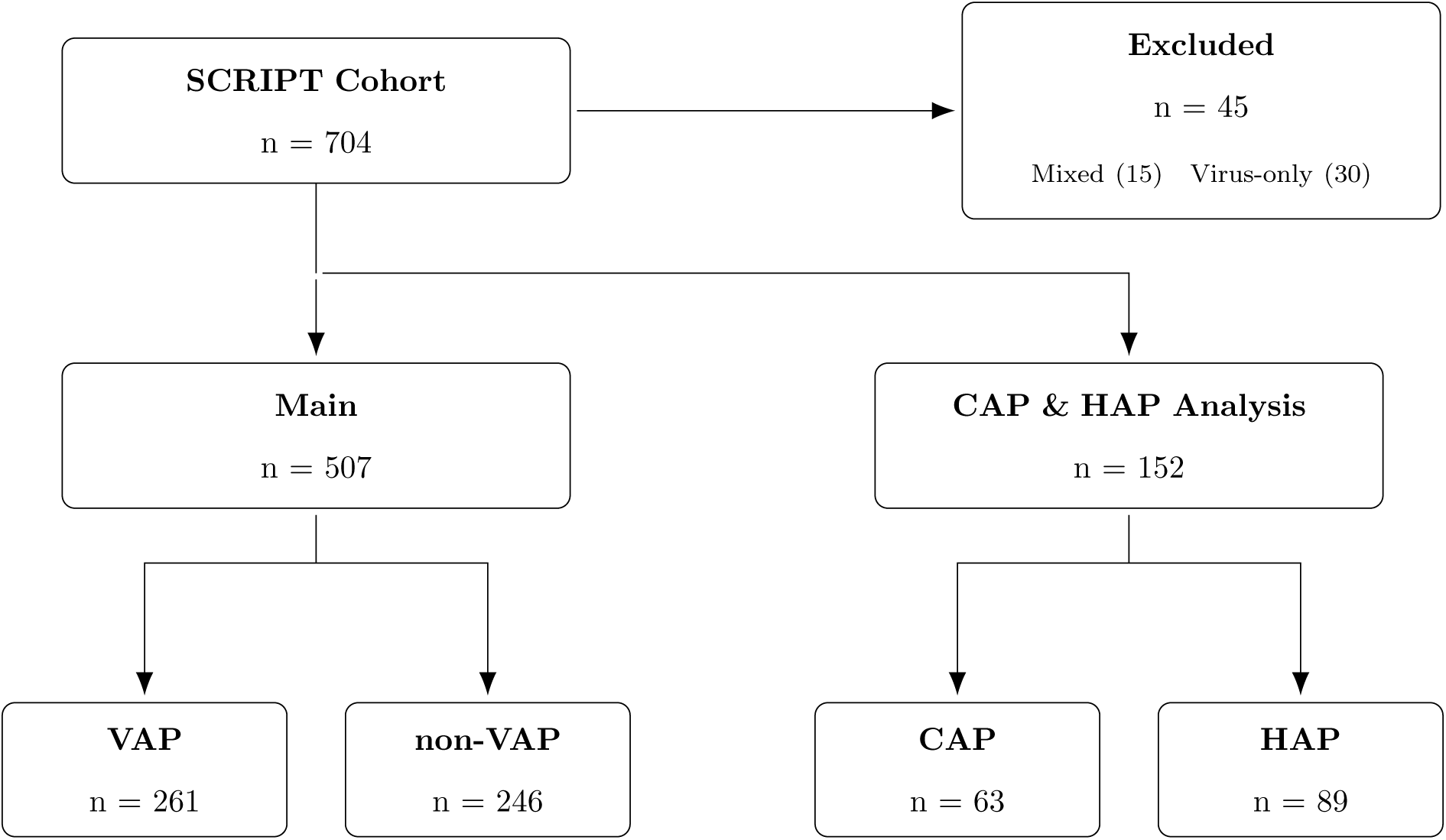
Patient flow diagram: cohort allocation to the primary VAP versus non-VAP analysis and the secondary CAP/HAP characterization. BAL-confirmed CAP/HAP episodes without superinfection or documented microbiologic cure were excluded from the primary analysis; CAP/HAP cases complicated by superinfection were classified as VAP, and successfully treated CAP/HAP cases were classified as non-VAP.

**Supplementary Table S1:** Clinical features from the SCRIPT cohort used for modeling during (7) event windows averaged, stratified by VAP status.

| Characteristic | Missing | Overall | Did not have VAP | Had VAP | P-Value |
| --- | --- | --- | --- | --- | --- |
| n |  | 4451 | 2040 | 2411 |  |
| Days on Ventilator | 0 | 8.0 [4.0,16.0] | 5.0 [3.0,10.0] | 11.0 [5.0,21.0] | <0.001 |
| Temperature (°F) | 3 | 98.6 [98.0,99.4] | 98.5 [98.0,99.2] | 98.7 [98.1,99.5] | <0.001 |
| Heart Rate (bpm) | 0 | 88.5 [76.9,100.7] | 85.9 [74.5,97.3] | 90.8 [79.1,102.9] | <0.001 |
| Mean Arterial Pressure (mmHg) | 8 | 77.6 [72.5,86.0] | 78.6 [72.9,87.5] | 77.0 [72.4,84.7] | <0.001 |
| Respiratory Rate (breaths/min) | 29 | 22.6 [18.8,27.0] | 21.1 [18.0,25.2] | 24.0 [20.0,28.0] | <0.001 |
| Oxygen Saturation (%) | 0 | 96.0 [94.5,97.5] | 96.4 [94.9,97.8] | 95.6 [94.1,97.0] | <0.001 |
| GCS Eye Opening | 52 | 3.0 [1.7,4.0] | 3.2 [2.2,4.0] | 2.7 [1.0,3.8] | <0.001 |
| GCS Motor Response | 52 | 4.7 [2.2,6.0] | 5.2 [4.0,6.0] | 4.0 [1.0,5.8] | <0.001 |
| RASS Score | 57 | -1.8 [-3.5,-0.5] | -1.3 [-2.7,-0.4] | -2.2 [-4.0,-0.7] | <0.001 |
| PEEP (cmH<sub>2</sub>O) | 27 | 6.5 [5.0,10.0] | 5.0 [5.0,8.0] | 8.0 [5.0,12.0] | <0.001 |
| FiO<sub>2</sub> (%) | 0 | 40.0 [30.0,50.0] | 37.7 [30.0,46.2] | 40.0 [33.4,50.0] | <0.001 |
| Plateau Pressure (cmH<sub>2</sub>O) | 301 | 22.8 [18.0,28.0] | 20.2 [17.0,24.8] | 25.0 [20.0,29.8] | <0.001 |
| Minute Ventilation (L/min) | 14 | 9.1 [7.2,11.3] | 8.6 [6.9,10.4] | 9.8 [7.6,11.8] | <0.001 |
| Arterial pH | 1042 | 7.4 [7.4,7.5] | 7.4 [7.4,7.5] | 7.4 [7.4,7.4] | 0.359 |
| PaCO<sub>2</sub> (mmHg) | 1042 | 42.3 [36.2,50.3] | 39.0 [33.9,45.5] | 45.4 [38.8,52.8] | <0.001 |
| PaO<sub>2</sub> (mmHg) | 1043 | 93.2 [79.5,115.2] | 99.5 [84.0,123.0] | 89.0 [77.5,108.7] | <0.001 |
| WBC Count (10<sup>3</sup>/μL) | 64 | 11.2 [7.9,15.6] | 10.5 [7.2,15.5] | 11.7 [8.7,15.7] | <0.001 |
| Absolute Lymphocytes (cells/μL) | 1469 | 0.9 [0.5,1.4] | 0.8 [0.4,1.3] | 0.9 [0.6,1.5] | <0.001 |
| Absolute Neutrophils (cells/μL) | 1450 | 8.7 [6.0,13.0] | 8.4 [5.3,13.9] | 8.9 [6.3,12.7] | 0.059 |
| Hemoglobin (g/dL) | 39 | 8.4 [7.7,9.8] | 8.4 [7.6,9.9] | 8.4 [7.7,9.7] | 0.441 |
| Platelet Count (10<sup>3</sup>/μL) | 39 | 183.0 [102.0,274.0] | 162.0 [77.0,247.0] | 201.0 [120.0,294.0] | <0.001 |
| Bicarbonate (mEq/L) | 39 | 26.0 [23.0,31.0] | 25.0 [22.0,28.7] | 27.3 [24.0,32.0] | <0.001 |
| Creatinine (mg/dL) | 68 | 0.9 [0.6,1.7] | 1.1 [0.6,1.9] | 0.8 [0.5,1.5] | <0.001 |
| Albumin (g/dL) | 1193 | 2.7 [2.4,3.0] | 2.8 [2.4,3.1] | 2.7 [2.4,3.0] | 0.001 |
| Total Bilirubin (mg/dL) | 1247 | 0.8 [0.5,1.5] | 0.9 [0.6,2.0] | 0.7 [0.5,1.2] | <0.001 |
| C-Reactive Protein (mg/L) | 3547 | 145.0 [73.8,246.4] | 170.1 [84.1,288.6] | 141.4 [69.9,231.0] | 0.003 |
| D-dimer (ng/mL) | 3142 | 2579.0 [842.0,6868.0] | 3225.3 [1078.0,8647.0] | 2196.0 [775.5,5929.0] | <0.001 |
| Ferritin (ng/mL) | 3686 | 857.1 [478.8,1412.0] | 1074.6 [548.6,1724.6] | 809.2 [461.0,1319.2] | <0.001 |
| Lactate Dehydrogenase (U/L) | 2943 | 419.5 [298.0,589.2] | 420.0 [273.0,656.6] | 419.0 [311.0,573.2] | 0.649 |
| Lactate (mmol/L) | 2315 | 1.4 [1.1,2.0] | 1.5 [1.1,2.2] | 1.3 [1.0,1.8] | <0.001 |
| Procalcitonin (ng/mL) | 3086 | 0.9 [0.3,2.9] | 1.3 [0.4,3.6] | 0.7 [0.2,2.4] | <0.001 |
| ECMO use, n (%) | 0 | 802 (18.0) | 267 (13.1) | 535 (22.2) | <0.001 |
| CRRT use, n (%) | 0 | 740 (16.6) | 358 (17.5) | 382 (15.8) | 0.138 |
| Vasopressor Support, n (%) | 0 | 1982 (44.5) | 856 (42.0) | 1126 (46.7) | 0.002 |

**Supplementary Table S2:** Bacterial Pathogens detected by BAL culture and Biofire PCR in the SCRIPT cohort.

| Pathogen | Culture (n) | PCR (n) |
| --- | --- | --- |
| *Pseudomonas aeruginosa* | 55 | 49 |
| *Staphylococcus aureus* | 53 | 62 |
| *Enterococcus faecalis* | 33 | — |
| *Staphylococcus coagulase negative* | 24 | — |
| *Klebsiella pneumoniae* | 22 | — |
| *Klebsiella pneumoniae group* | — | 18 |
| *Escherichia coli* | 21 | 22 |
| Yeast, not *Cryptococcus* species | 21 | — |
| *Viridans streptococcus* | 17 | — |
| *Corynebacterium species* | 17 | — |
| *Enterobacter aerogenes* | 16 | — |
| *Enterococcus faecium* | 10 | — |
| *Citrobacter koseri* | 9 | — |
| *Stenotrophomonas maltophilia* | 8 | — |
| *Serratia marcescens* | 8 | 10 |
| *Enterobacter cloacae complex* | 8 | 11 |
| *Klebsiella oxytoca* | 6 | 5 |
| *Viridans Streptococcus* | 6 | — |
| *Acinetobacter baumannii* | 5 | — |
| *Acinetobacter calcoaceticus–baumannii complex* | — | 4 |
| *Lactobacillus species* | 4 | — |
| *Achromobacter xylosoxidans* | 4 | — |
| *Burkholderia cepacia complex* | 4 | — |
| *Beta-hemolytic streptococci, Group F* | 4 | — |
| *Hafnia alvei* | 3 | — |
| *Streptococcus agalactiae, Group B* | 2 | 3 |
| *Proteus mirabilis* | 2 | — |
| *Proteus spp.* | — | 3 |
| *Granulicatella species* | 2 | — |
| *Chryseobacterium indologenes* | 2 | — |
| *Beta-hemolytic streptococci, Group G* | 1 | — |
| *Pseudomonas fluorescens* | 1 | — |
| *Enterobacter cloacae* | 1 | — |
| *Elizabethkingia meningoseptica* | 1 | — |
| *Candida auris* | 1 | — |
| *Klebsiella aerogenes* | 1 | 18 |
| *Enterococcus avium* | 1 | — |
| *Enterococcus raffinosus* | 1 | — |
| *Granulicatella adiacens* | 1 | — |
| *Candida albicans* | 1 | — |
| *Chryseobacterium gleum* | 1 | — |
| *Staphylococcus epidermidis* | 1 | — |
| *Streptococcus pseudopneumoniae* | 1 | — |
| *Haemophilus influenzae* | 1 | 7 |
| *Burkholderia gladioli* | 1 | — |
| *Citrobacter freundii group* | 1 | — |
| *Neisseria species* | 1 | — |
| *Achromobacter denitrificans* | 1 | — |
| *Beta-hemolytic streptococci, Group C* | 1 | — |
| *Enterococcus species* | 1 | — |
| *Staphylococcus intermedius* | 1 | — |
| *Streptococcus pneumoniae* | — | 2 |
| *Streptococcus pyogenes* | — | 1 |

**Supplementary Figure S2:**
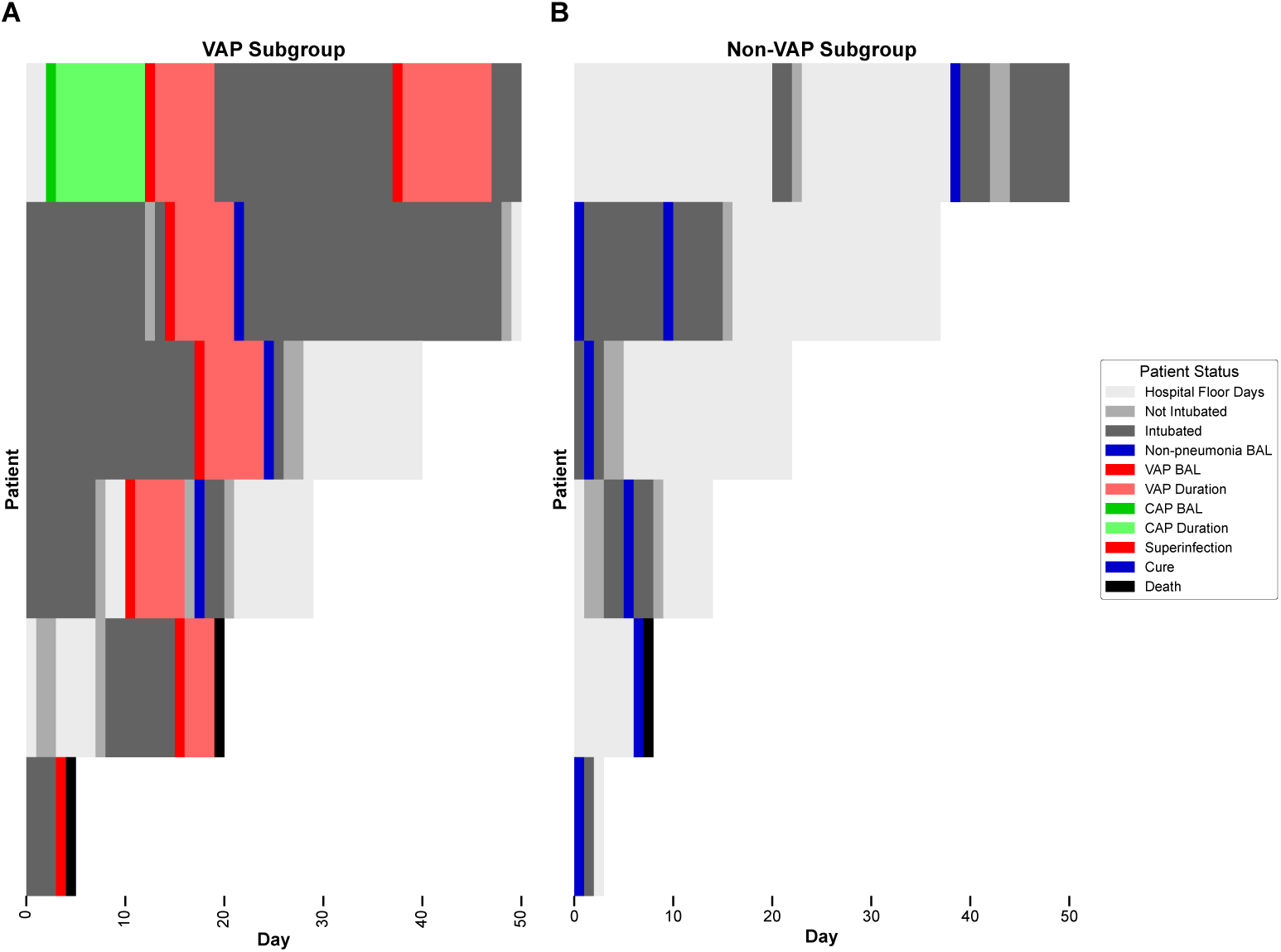
Patient-level timelines illustrating pneumonia adjudication and clinical trajectories. Selected patient hospital stays highlight temporal relationships between intubation, bronchoalveolar lavage (BAL) events, and clinical outcomes. Each row represents an individual patient plotted by hospital day. (A) Patients with BAL-confirmed VAP only, showing intubation periods (gray), VAP BAL-positive days (red), VAP episode duration (pink), and outcomes including cure (blue) and death (black). (B) Patients with BAL-confirmed non-VAP status only, with BAL-negative events marked in blue and corresponding intubation periods shaded. This zoomed view emphasizes the heterogeneity in timing, duration, and outcomes of pneumonia-related events across individual patient trajectories.

**Supplementary Figure S3:**
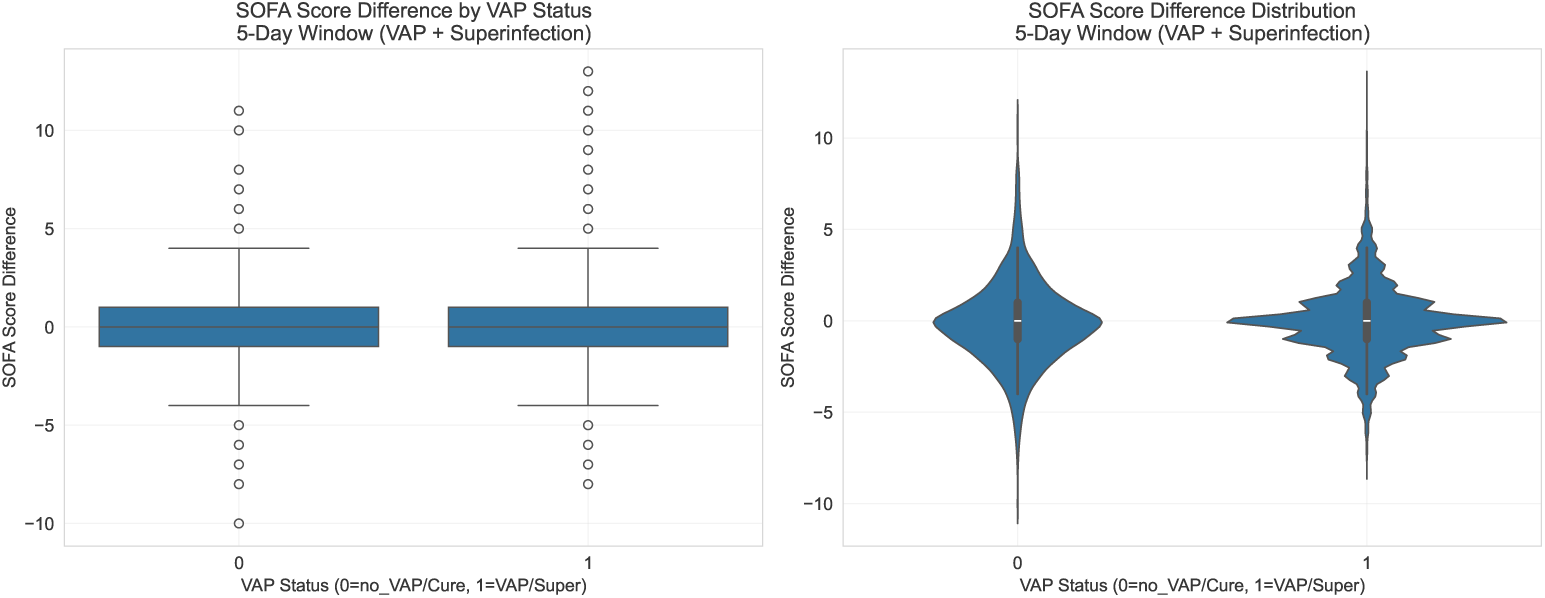
Distribution of SOFA score differences between the first and second intubated days, stratified by VAP status.

**Supplementary Figure S4:**
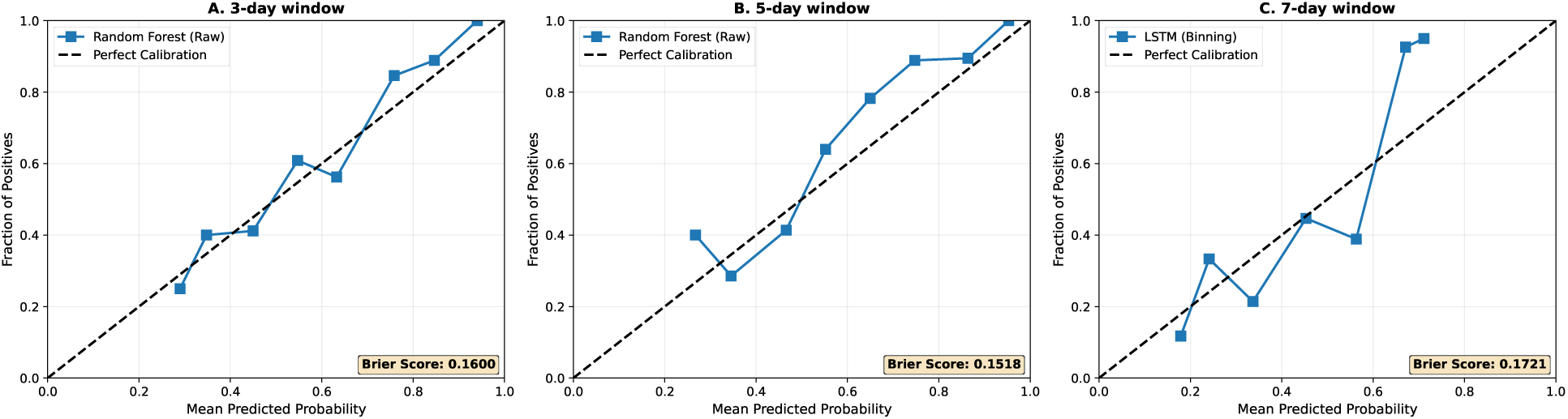
Calibration performance of top models across prediction windows. Panels A–C show calibration curves for the best-performing model for the 3-, 5-, and 7-day VAP prediction tasks. Random Forest models were best calibrated for the 3- and 5-day windows, while an LSTM model performed best for the 7-day task. The dashed line indicates perfect calibration, and Brier scores are reported in each panel.

**Supplementary Figure S5:**
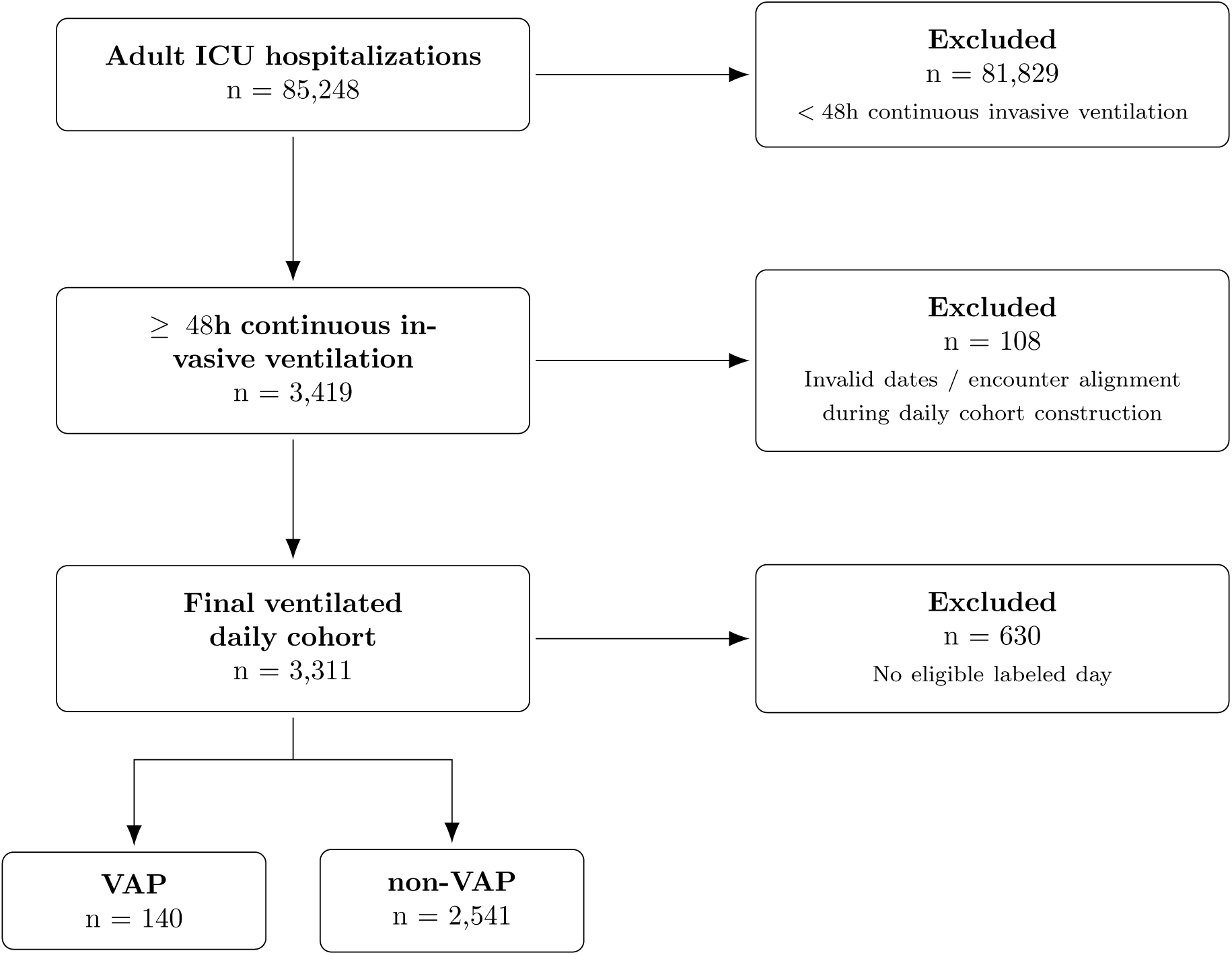
Patient flow of MIMIC-IV cohort selection and allocation to the primary analysis (VAP vs. non-VAP). Patients were filtered to those with *≥* 48 hours of continuous invasive ventilation, expanded into a daily cohort, and categorized by VAP or non-VAP. Patients without any eligible labeled day were excluded.

**Supplementary Figure S6:**
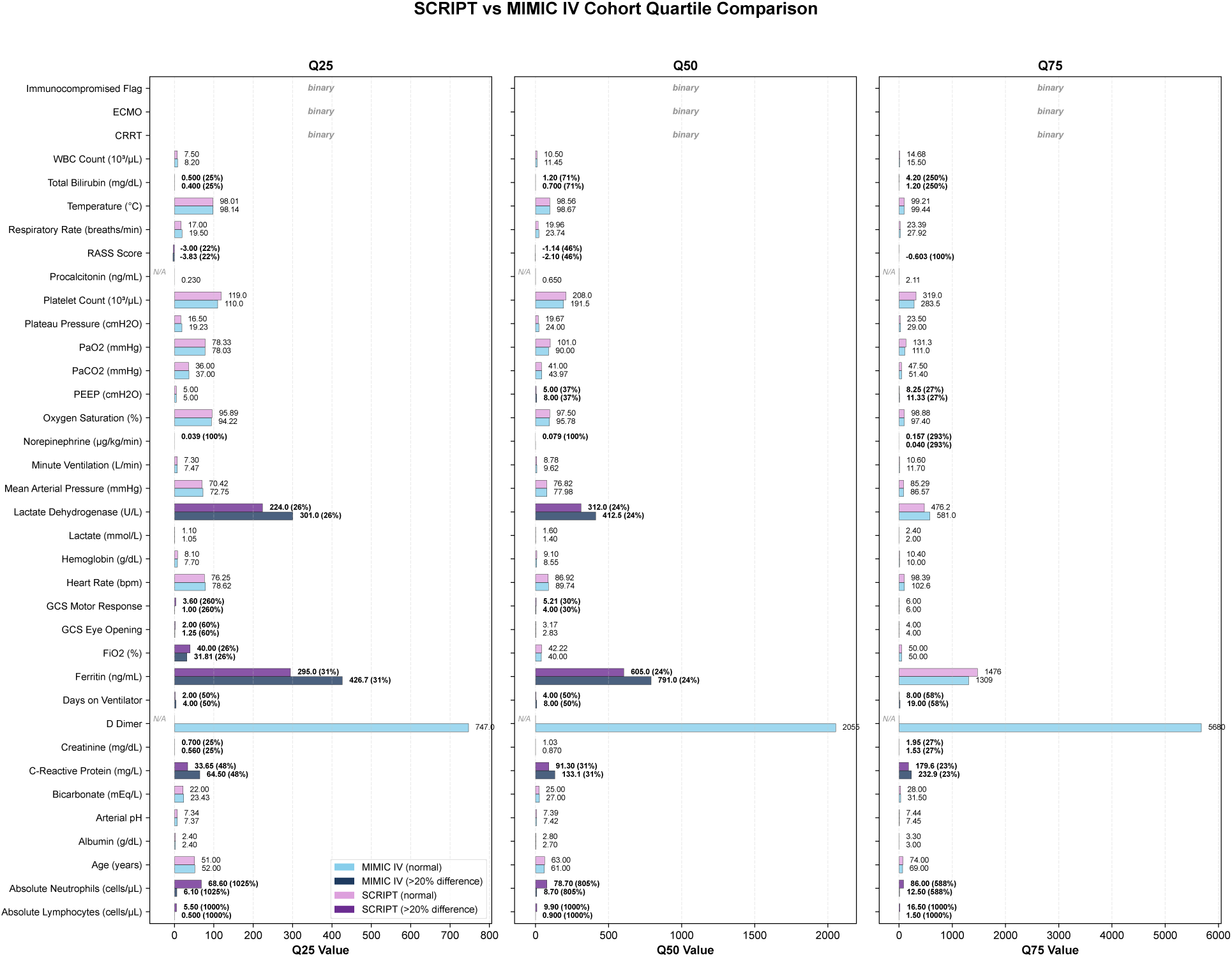
Comparison of feature distributions between SCRIPT and MIMIC IV cohorts using quartiles (Q25, Q50, Q75). Each panel displays the 25th percentile, median, and 75th percentile values for clinical features, with darker bars indicating *>* 20% difference between cohorts.

**Supplementary Figure S7:**
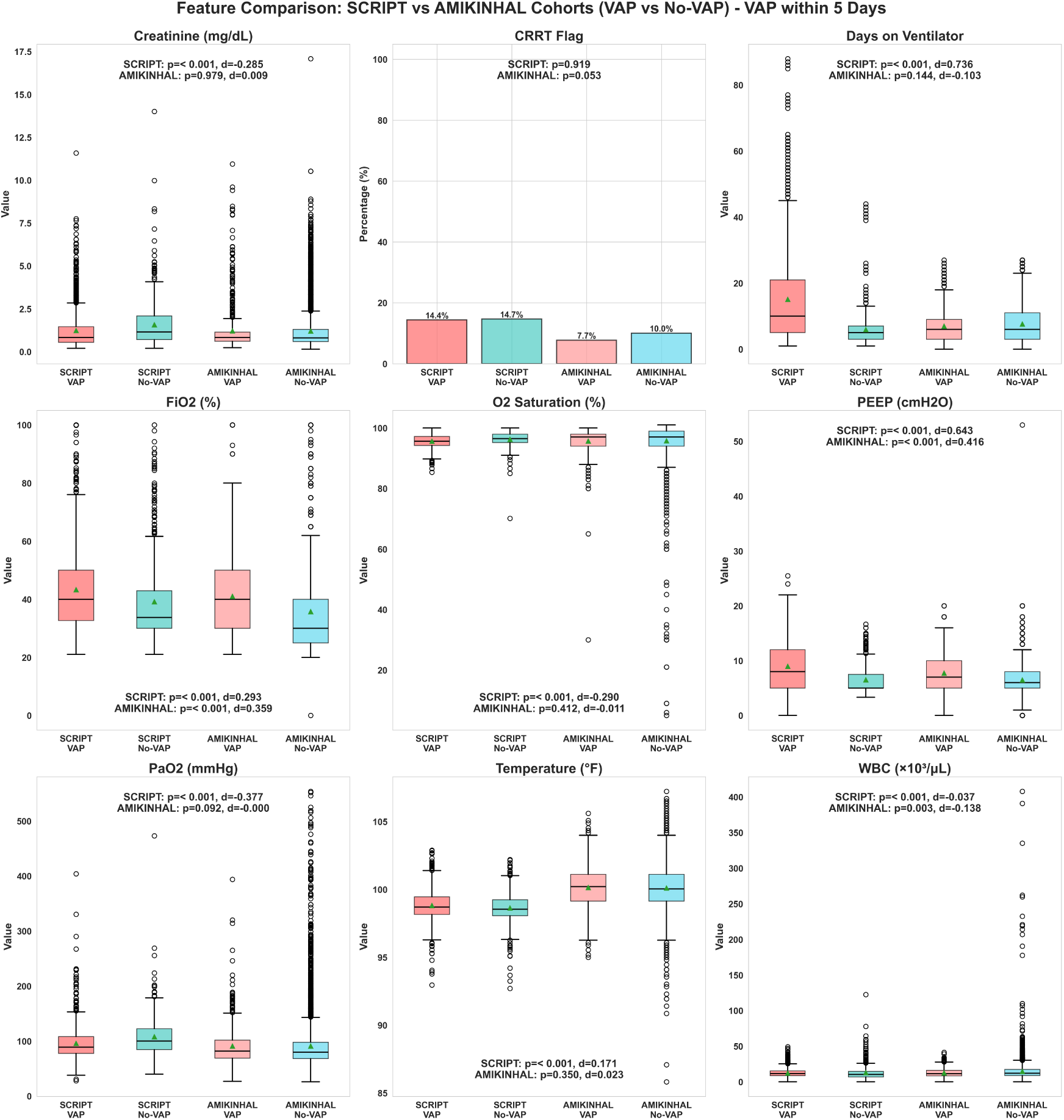
Feature comparison between SCRIPT and AMIKINHAL cohorts for VAP prediction within 5 days. Panels show distributions of clinical features comparing VAP (label=1) versus non-VAP (label=0) patient-days for both cohorts. P-values were calculated using Mann-Whitney U test for continuous variables and chi-square test for binary variables (CRRT flag).

**Supplementary Table S3:**
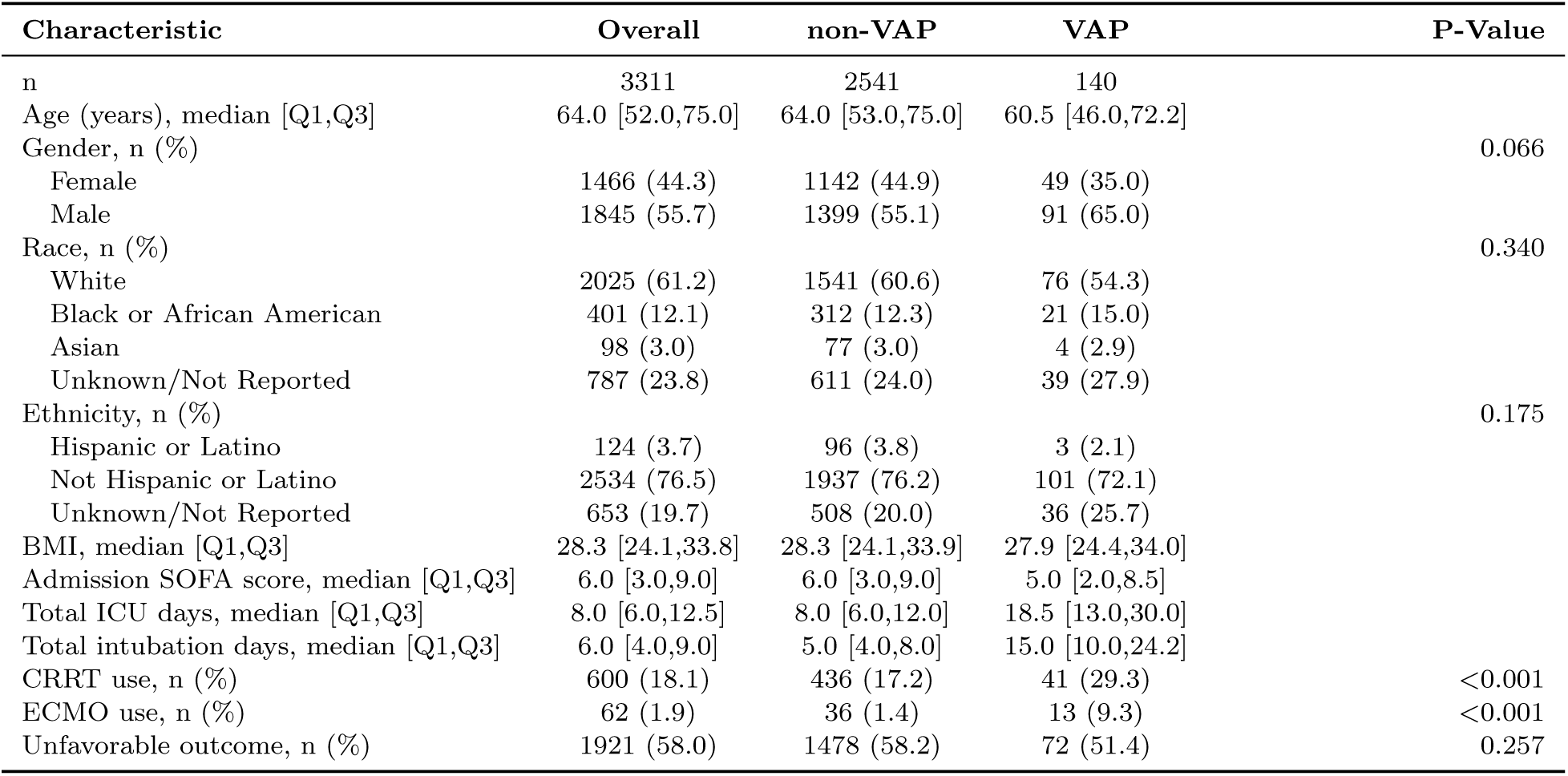
Baseline demographics and clinical characteristics in the MIMIC-IV cohort, stratified by VAP status. VAP status was defined using a first eligible labeled day approach, categorizing patients as VAP or non-VAP.

| Characteristic | Overall | non-VAP | VAP | P-Value |
| --- | --- | --- | --- | --- |
| n | 3311 | 2541 | 140 |  |
| Age (years), median [Q1,Q3] | 64.0 [52.0,75.0] | 64.0 [53.0,75.0] | 60.5 [46.0,72.2] |  |
| Gender, n (%) |  |  |  | 0.066 |
| Female | 1466 (44.3) | 1142 (44.9) | 49 (35.0) |  |
| Male | 1845 (55.7) | 1399 (55.1) | 91 (65.0) |  |
| Race, n (%) |  |  |  | 0.340 |
| White | 2025 (61.2) | 1541 (60.6) | 76 (54.3) |  |
| Black or African American | 401 (12.1) | 312 (12.3) | 21 (15.0) |  |
| Asian | 98 (3.0) | 77 (3.0) | 4 (2.9) |  |
| Unknown/Not Reported | 787 (23.8) | 611 (24.0) | 39 (27.9) |  |
| Ethnicity, n (%) |  |  |  | 0.175 |
| Hispanic or Latino | 124 (3.7) | 96 (3.8) | 3 (2.1) |  |
| Not Hispanic or Latino | 2534 (76.5) | 1937 (76.2) | 101 (72.1) |  |
| Unknown/Not Reported | 653 (19.7) | 508 (20.0) | 36 (25.7) |  |
| BMI, median [Q1,Q3] | 28.3 [24.1,33.8] | 28.3 [24.1,33.9] | 27.9 [24.4,34.0] |  |
| Admission SOFA score, median [Q1,Q3] | 6.0 [3.0,9.0] | 6.0 [3.0,9.0] | 5.0 [2.0,8.5] |  |
| Total ICU days, median [Q1,Q3] | 8.0 [6.0,12.5] | 8.0 [6.0,12.0] | 18.5 [13.0,30.0] |  |
| Total intubation days, median [Q1,Q3] | 6.0 [4.0,9.0] | 5.0 [4.0,8.0] | 15.0 [10.0,24.2] |  |
| CRRT use, n (%) | 600 (18.1) | 436 (17.2) | 41 (29.3) | <0.001 |
| ECMO use, n (%) | 62 (1.9) | 36 (1.4) | 13 (9.3) | <0.001 |
| Unfavorable outcome, n (%) | 1921 (58.0) | 1478 (58.2) | 72 (51.4) | 0.257 |

**Supplementary Table S4:** Clinical features in the MIMIC-IV cohort used for modeling during event windows, stratified by VAP status.

| Characteristic | Missing | Overall | Did not have VAP | Had VAP | P-Value |
| --- | --- | --- | --- | --- | --- |
| n |  | 6667 | 5848 | 819 |  |
| Days on Ventilator | 0 | 2.0 [1.0,2.0] | 2.0 [1.0,2.0] | 5.0 [3.0,12.0] | <0.001 |
| Temperature (°F) | 512 | 98.6 (1.3) | 98.5 (1.3) | 99.0 (1.1) | <0.001 |
| Heart Rate (bpm) | 3 | 87.3 (17.4) | 87.3 (17.6) | 87.7 (15.7) | 0.444 |
| Mean Arterial Pressure (mmHg) | 4 | 77.2 (10.5) | 77.1 (10.7) | 77.4 (9.4) | 0.482 |
| Respiratory Rate (breaths/min) | 8 | 19.9 (4.7) | 19.7 (4.5) | 21.3 (5.6) | <0.001 |
| Oxygen Saturation (%) | 13 | 97.6 (2.2) | 97.7 (2.3) | 97.2 (2.1) | <0.001 |
| GCS Eye Opening | 105 | 2.2 (1.0) | 2.2 (1.0) | 2.1 (1.1) | 0.088 |
| GCS Motor Response | 115 | 3.8 (1.8) | 3.8 (1.8) | 3.5 (2.0) | <0.001 |
| RASS Score | 869 | -2.6 (1.7) | -2.6 (1.7) | -2.6 (1.9) | 0.411 |
| PEEP (cmH<sub>2</sub>O) | 5 | 7.0 (3.1) | 6.8 (2.9) | 8.5 (3.5) | <0.001 |
| FiO<sub>2</sub> (%) | 1 | 51.6 (15.7) | 51.8 (15.9) | 50.2 (14.1) | 0.003 |
| Plateau Pressure (cmH<sub>2</sub>O) | 1177 | 19.7 (4.9) | 19.4 (4.8) | 22.1 (5.2) | <0.001 |
| Minute Ventilation (L/min) | 9 | 9.1 (2.5) | 9.0 (2.4) | 9.7 (3.0) | <0.001 |
| Arterial pH | 816 | 7.4 (0.1) | 7.4 (0.1) | 7.4 (0.1) | <0.001 |
| PaCO<sub>2</sub> (mmHg) | 840 | 42.3 (10.0) | 42.0 (10.0) | 44.5 (9.6) | <0.001 |
| PaO<sub>2</sub> (mmHg) | 840 | 125.7 (67.7) | 127.3 (69.9) | 114.3 (47.1) | <0.001 |
| WBC Count (10<sup>3</sup>/μL) | 414 | 14.0 (9.7) | 14.1 (10.0) | 13.8 (7.6) | 0.429 |
| Absolute Lymphocytes (cells/μL) | 5835 | 1.5 (2.8) | 1.4 (2.8) | 1.9 (3.4) | 0.338 |
| Absolute Neutrophils (cells/μL) | 5848 | 12.8 (8.0) | 12.7 (8.0) | 14.0 (8.4) | 0.236 |
| Hemoglobin (g/dL) | 396 | 10.0 (2.0) | 10.1 (2.1) | 9.6 (1.8) | <0.001 |
| Platelet Count (10<sup>3</sup>/μL) | 415 | 193.2 (117.5) | 194.1 (118.3) | 186.5 (111.7) | 0.074 |
| Bicarbonate (mEq/L) | 391 | 22.9 (5.2) | 22.5 (5.1) | 25.2 (4.9) | <0.001 |
| Creatinine (mg/dL) | 386 | 1.7 (1.6) | 1.8 (1.6) | 1.5 (1.2) | <0.001 |
| Albumin (g/dL) | 4614 | 2.8 (0.7) | 2.8 (0.7) | 2.7 (0.7) | 0.012 |
| Total Bilirubin (mg/dL) | 3192 | 3.1 (6.3) | 3.2 (6.6) | 2.6 (3.8) | 0.012 |
| C-Reactive Protein (mg/L) | 6529 | 157.7 (105.7) | 156.4 (106.5) | 169.1 (101.9) | 0.666 |
| Ferritin (ng/mL) | 6426 | 3796.5 (9368.5) | 3943.5 (9657.1) | 1730.2 (2599.4) | 0.019 |
| Lactate Dehydrogenase (U/L) | 4534 | 745.7 (1217.0) | 757.8 (1260.3) | 643.2 (752.9) | 0.048 |
| Lactate (mmol/L) | 1796 | 2.6 (2.2) | 2.7 (2.2) | 1.9 (1.4) | <0.001 |
| ECMO use, n (%) | 0 | 147 (2.2) | 76 (1.3) | 71 (8.7) | <0.001 |
| CRRT use, n (%) | 0 | 609 (9.1) | 450 (7.7) | 159 (19.4) | <0.001 |

**Supplementary Figure S8:**
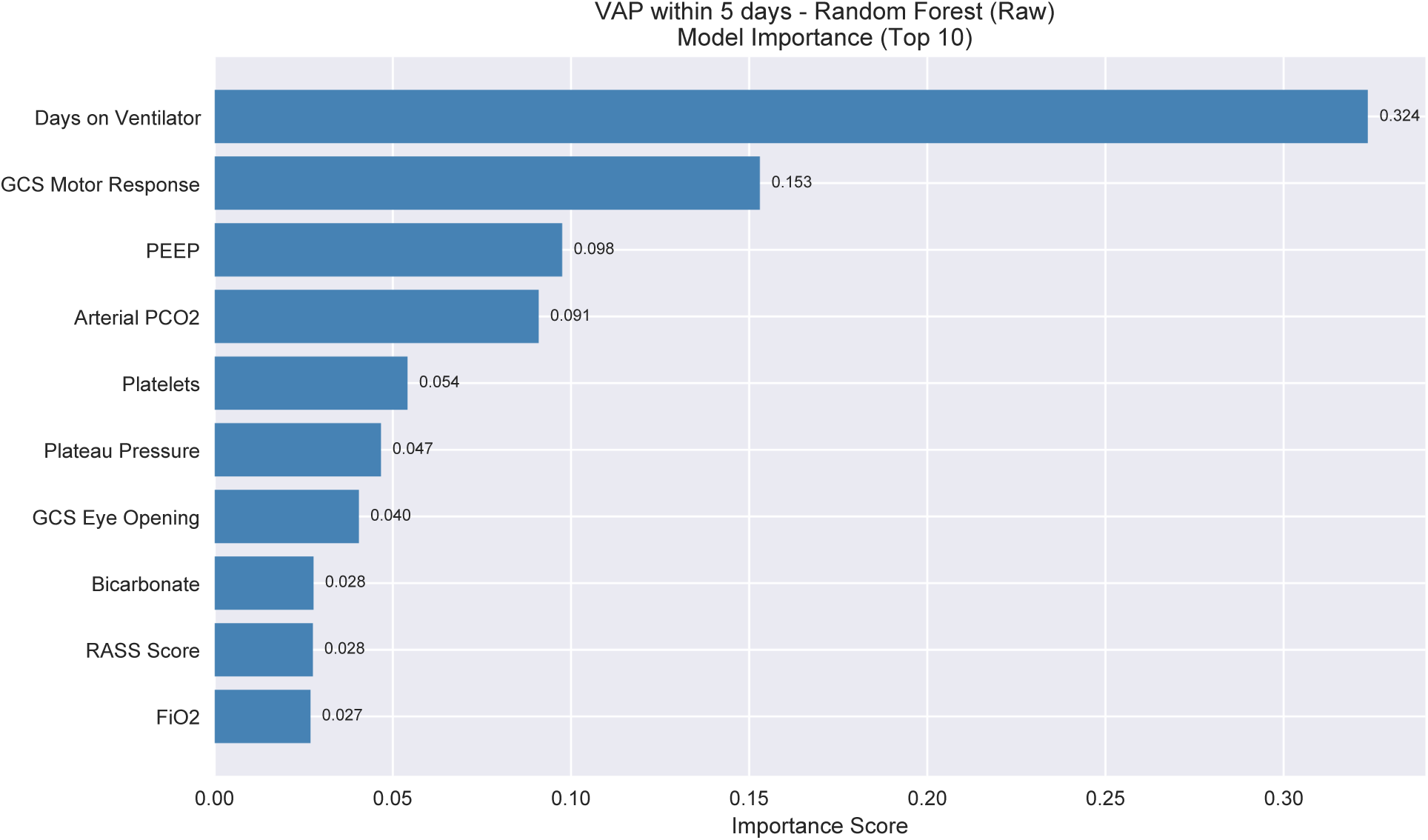
Feature importance for VAP prediction within 5 days in the external MIMIC-IV cohort. Impurity-based feature importance (model importance) for the raw-feature Random Forest model corresponding to the best-performing external VAP within 5 days task. The most influential predictors include duration of mechanical ventilation, neurologic status, and ventilatory mechanics.

